# One Million and Counting: Estimates of Deaths in the United States from Ancestral SARS-CoV-2 and Variants

**DOI:** 10.1101/2022.05.31.22275835

**Authors:** Jo Walker, Nathan D. Grubaugh, Gregg Gonsalves, Virginia Pitzer, Zain Rizvi

## Abstract

**Background:** Over one million COVID-19 deaths have been recorded in the United States. Sustained global SARS-CoV-2 transmission has led to the emergence of new variants with increased transmissibility, virulence, and/or immune evasion. The specific burden of mortality from each variant over the course of the U.S. COVID-19 epidemic remains unclear.

**Methods:** We constructed an epidemiologic model using data reported by the CDC on COVID-19 mortality and circulating variant proportions to estimate the number of recorded COVID-19 deaths attributable to each SARS-CoV-2 variant in the U.S. We conducted sensitivity analysis to account for parameter uncertainty.

**Findings:** Of the 1,003,419 COVID-19 deaths recorded as of May 12, 2022, we estimate that 460,124 (46%) were attributable to WHO-designated variants. By U.S. Census Region, the South recorded the most variant deaths per capita (median estimate 158 per 100,000), while the Northeast recorded the fewest (111 per 100,000). Over 40 percent of national COVID-19 deaths were estimated to be caused by the combination of Alpha (median estimate 39,548 deaths), Delta (273,801), and Omicron (117,560).

**Interpretation:** SARS-CoV-2 variants that have emerged around the world have imposed a significant mortality burden in the U.S. In addition to national public health strategies, greater efforts are needed to lower the risk of new variants emerging, including through global COVID-19 vaccination, treatment, and outbreak mitigation.

## Introduction

Over one million deaths have been recorded in the United States from coronavirus disease 2019 (COVID-19). Since severe acute respiratory syndrome coronavirus 2 (SARS-CoV-2) emerged in Wuhan, China in late 2019, the virus has continued to evolve, with multiple new variants characterized by increased transmissibility, virulence, and/or immune evasion.

Variants reported around the world have repeatedly changed the trajectory of the U.S. epidemic. As of May 12, 2022, the World Health Organization (WHO) has named five variants of concern (VOC) that were originally detected in four different continents [**Table 1**]. All five variants of concern were more transmissible than the original virus, and spread to the U.S.^1–3^ Delta (lineage B.1.617.2) was particularly severe from a clinical standpoint, and drove a surge of hospitalizations across the U.S in the summer and fall of 2021.^4^ While Omicron (lineage B.1.1.529) appears to cause less severe disease than Delta on a per-infection basis—likely due to a combination of lower intrinsic severity and the high observed concentration of cases among the previously vaccinated or infected^5^—its elevated transmissibility, including among the seropositive, resulted in the largest wave of SARS-CoV-2 transmission in the U.S. to date.^6, 7^ Vaccines developed against the non-variant SARS-CoV-2 are less effective at protecting against infection with Delta and Omicron, but remain highly effective against severe disease.^8–10^

**Table 1.** SARS-CoV-2 variants of concern designated by the World Health Organization.^11, 12^

| **Pango Lineage** | **WHO Designation** | **First Detection Location** | **Earliest Documented Samples** | **Earliest U.S. Report** | **Downgrade in Status** |
| --- | --- | --- | --- | --- | --- |
| B.1.1.7 | Alpha | United Kingdom | Sept. 2020 | Dec. 2020 | Previous variant of concern: Mar. 2022 |
| B.1.351 | Beta | South Africa | May 2020 | Jan. 2021 | Previous variant of concern: Mar. 2022 |
| P.1 | Gamma | Brazil | Nov. 2020 | Jan. 2021 | Previous variant of concern: Mar. 2022 |
| B.1.617.2 | Delta | India | Oct. 2020 | Mar. 2021 | Not downgraded as of May 2022 |
| B.1.1.529 | Omicron | Botswana and South Africa | Nov. 2021 | Dec. 2021 | Not downgraded as of May 2022 |

In addition to variants of concern, the WHO and U.S. Centers for Disease Control and Prevention (CDC) have classified a number of additional SARS-CoV-2 lineages as variants of interest (VOI) or under monitoring (VUM).^13^ The variants have emerged in the context of sustained global transmission. More than 500 million COVID-19 cases have been reported globally, reflecting only a fraction of total infections, and giving SARS-CoV-2 many opportunities to mutate and evolve.^14^ One third of the global population has still not received a vaccine dose, limiting the benefits of vaccination on reducing transmission and viral replication at a population level.^15^

Recent studies have described clinical outcomes associated with SARS-CoV-2 variants, and analyzed factors associated with COVID-19 mortality in the U.S. including race and geography.^16, 17^ The aim of our study is to estimate the burden of deaths associated with each SARS-CoV-2 variant in the U.S. We construct a model using data reported by the CDC on COVID-19 deaths by jurisdiction and circulating variant proportions.

## Methods

To estimate the number of recorded COVID-19 deaths attributable to each variant, we back-distributed deaths to estimate the number of ultimately fatal COVID-19 cases testing positive on each day in each jurisdiction, and compared these counts to proportions of variants among sequenced cases in the same time period and location, adjusted for differences in disease severity between variants.

### Data

Our analysis relies on provisional counts of COVID-19 deaths from the CDC’s National Center for Health Statistics (NCHS).^18^ This dataset is based on death certificate records and reports the number of deaths with confirmed or presumed COVID-19 by jurisdiction (the 50 states, New York City, District of Columbia, and Puerto Rico) and time period (week, month, year, and total) of occurence. All data were accessed May 12, 2022.

The NCHS data are subject to change as new death certificate records are received and processed by the CDC. While the majority of COVID-19 deaths are reported within 10 days of the date of death, state reporting timelines vary significantly.^19^ Counts from recent weeks may be incomplete. However, because adjustments are almost always upwards, the NCHS provisional COVID-19 deaths can be considered a lower bound of the number of recorded COVID-19 deaths that have occurred.

NCHS death counts are also displayed by the week of occurrence, rather than the day of occurrence. Since the delay distributions that we applied to match deaths to the appropriate variant proportions (see below) require analysis at the level of days, we imputed daily deaths within each week by sampling from a uniform distribution. Finally, for each jurisdiction, the NCHS suppresses death counts between 1 and 9 for privacy reasons. As a result, the specific week is not known for 3,628 of the 1,003,419 COVID-19 deaths recorded in this dataset (0.4% of deaths overall, 0.002% to 39.4% of deaths in each jurisdiction). In this analysis, we do not assign these deaths to a week or otherwise attribute them to individual variants, meaning that our variant-specific death estimates should be treated as lower bounds.

Variant proportions were retrieved either directly^20^ from the CDC, or based on code and data made publicly available^21^ by the CDC. We considered 26 mutually-exclusive categories that deaths could be assigned to: the 5 current and former variants of concern (Alpha, Beta, Gamma, Delta, and Omicron), 19 other variants and lineages which have been recognized by the WHO and detected in the U.S. (A.23.1, A.27, A.28, B.1.1.318, B.1.1.519, B.1.214.2, B.1.617.3, B.1.620, B.1.640, C.16, C.36, Epsilon, Eta, Iota, Kappa, Lambda, Mu, Theta, or Zeta), a composite group of the early non-VOI/VOC/VBM virus lineages, which we will refer to as “non-variant”, and “unknown” (deaths for which the week of occurrence is suppressed).^9^

CDC genomic surveillance data was available from January 3rd, 2021. For the purpose of this analysis, we assumed that no variants were present in the U.S. prior to this date. In reality, there is evidence for circulation of the Alpha^2, 22, 23^ Epsilon^24^, and Iota^22^ variants in the U.S. in late 2020, albeit at low frequencies: thus, estimates of the mortality burden for these variants should be considered conservative. For the period between January 3rd, 2021, and January 1st, 2022, we used publicly available^19^ code and data underpinning the CDC’s variant proportion analysis to generate weighted variant proportions for each jurisdiction and U.S. Health and Human Services (HHS) region in non-overlapping weekly intervals. If no data on sequenced cases was available for a week in any given jurisdiction, we substituted variant proportions from the wider HHS region.

For the period after January 1st, 2022, the code and data described above was not publicly available. Instead, we used variant proportions retrieved directly from the CDC for national and HHS-regional levels in non-overlapping two-week intervals.^18^ We assumed that all states in a region had the same variant distribution during any given 2-week period. This simplifying assumption likely only had a marginal impact on our results, since the overwhelming majority of cases sampled in this period were attributed to Omicron.

### Analysis

Given the delays between symptom onset, testing, and death, we applied lags to infer the likely time of sample collection for new deaths. For our primary analysis, we assumed that sample collection for testing always took place either 0, 1, 2, or 3 days after symptom onset, with an associated probability of 25% for each of these 4 possible lag times. This is a simplifying assumption on our part. We also assumed the lag from symptom onset to death, *lag*_*onset to death*_, follows a lognormal distribution with a mean and standard deviation of 20.2 and 11.6 days, respectively (mean and standard deviation of the log(lag) = 2.863 and 0.534 respectively), truncated at 50 days. This corresponds to the best-fitting distribution for right-censored onset-to-death observations in Linton et al.^25^

Before applying variant proportions to COVID-19 deaths, we adjusted variant proportions among cases for variant-specific differences in disease severity. For instance, during periods where Omicron and Delta co-circulated, we expect Delta to be overrepresented among deaths relative to its share of cases (most of which will not be fatal), since Delta infections are more likely to result in severe disease than Omicron infections (whether due to higher intrinsic severity, a greater concentration of cases in unvaccinated individuals, or a combination).^5^

We derived a variant *i*’s proportion of fatal cases, *p*_*i,fatal*_ from its share of all cases *p*_*i,all*_ and the relative risk of death *RR*_*i*_ among confirmed cases infected with variant *i* compared to non-variant virus in a given time period *t* as

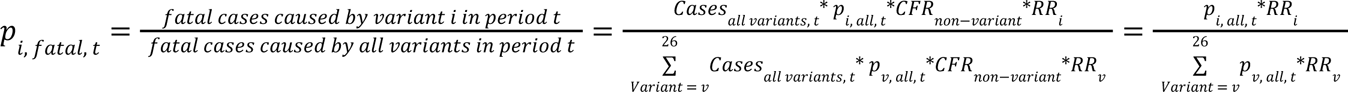

To parameterize *RR*_*i*_, we derived data on the relative severity of variants from a European Centre for Disease Prevention and Control (ECDC) review which was last updated in early April 2022.^26^ This review found no evidence for differences in severity (relative to the dominant variant(s) at the time of emergence) for the B.1.617.3, Epsilon, Eta, Iota, Kappa, Lambda, Mu, Theta, or Zeta variants. Evidence of differential severity was reported for the Alpha, Beta, Gamma, Delta, and Omicron variants [**Table 2**].

**Table 2.**
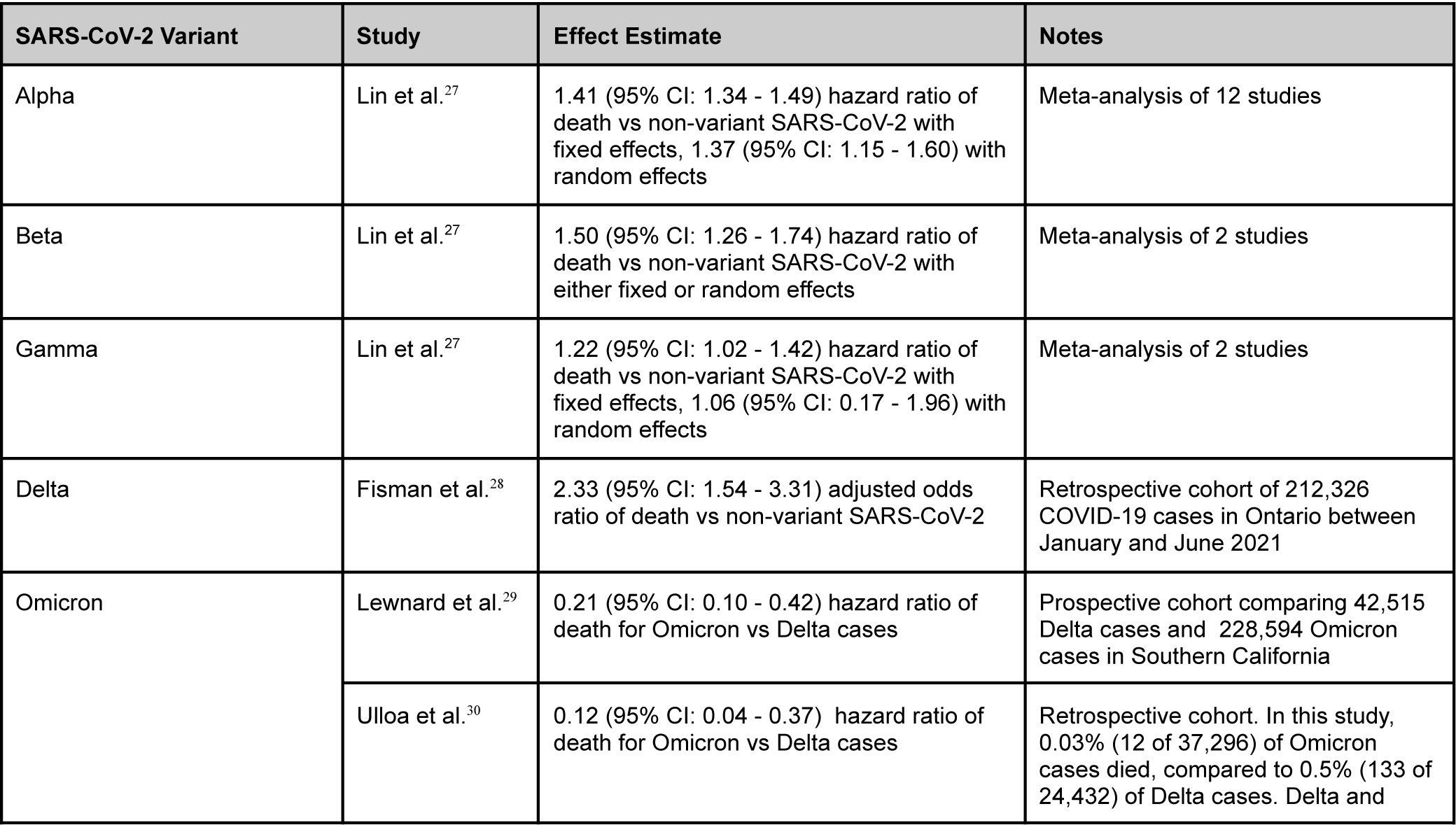
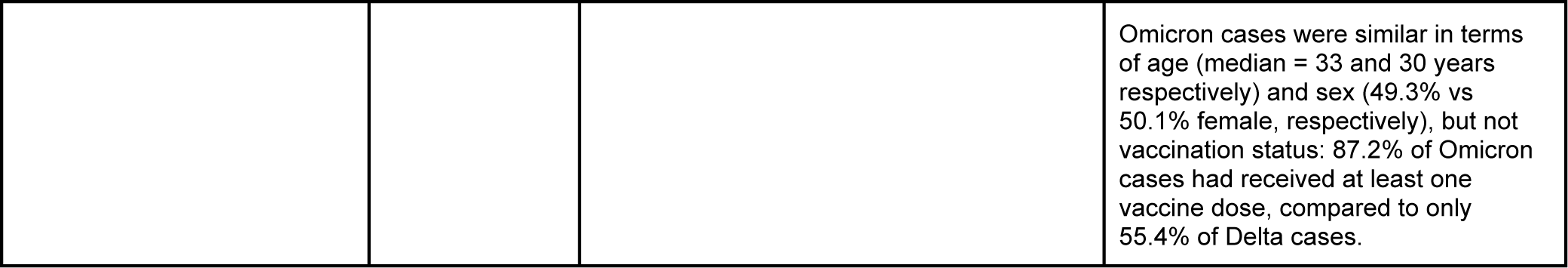
Estimates of the relative risk of death for different SARS-CoV-2 Variants of Concern

Based on the estimates above, we assumed that *RR*_*i*_ (the relative risk of death among confirmed cases caused by variant *i* vs non-variant in any given period of time), is 1.4 for Alpha, 1.5 for Beta, 1.2 for Gamma, 2.3 for Delta, 0.5 for Omicron (∼0.21\**RR*_*Delta*_), and 1 for the non-variant and all other variants in the primary analysis.

As a sensitivity analysis, we varied the assumed relative severity of Omicron from *RR*_*omicron*_ = 0. 15, the approximate ratio of observed Omicron and Delta case fatality rates in 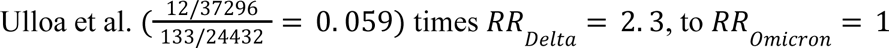, which corresponds approximately to the upper 95% bound of the Omicron vs Delta hazard ratio of death estimated by Lewnard et al., 0.42, times *RR_delta_* = 2. 3.^29, 30^ The results from these sensitivity analyses provide upper and lower limits for the plausible number of recorded COVID-19 deaths caused by the Omicron variant.

Finally, after deriving appropriate severity-adjusted variant proportion values *p*_*i, fatal*_ for each jurisdiction and time period, we estimated the number of recorded COVID-19 deaths caused by each variant *i* as

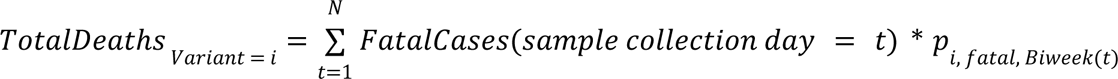

for each jurisdiction, where *Biweek*(*t*) species the non-overlapping two-week period containing day *t*. We reported national, regional (based on the four U.S. Census Regions.^31^ For this analysis, we included Puerto Rico with states in the *South* region), and jurisdiction-specific estimates.

Median estimates and 95% credible intervals were derived by running the model described above 250 times. Within each model run, we performed bootstrap resampling of the variant proportions at each location and time period before adjusting for severity, and resampled the day of death within the NCHS-specified week, the onset-to-death delay, and the onset-to-testing delay for each death in the dataset.

## Results

### National estimates

Of the 1,003,419 (299 per 100,000 people) recorded COVID-19 deaths in our dataset at the national level (the 50 states, the District of Columbia, and Puerto Rico), our median estimate is that 539,667 (95% credible interval: 538,928 - 540,466 deaths, 54% of all COVID-19 deaths, 161 deaths per 100,000) were caused by non-variant SARS-CoV-2, while 460,124 (95% CI: 459,325 - 460,863 deaths, 46% of deaths, 137 per 100,000) were caused by a WHO-classified variant [**Table 3**]. Of the variants that have had the greatest impact on the COVID-19 epidemic in the United States, we estimate that the Alpha variant caused a median of 39,548 recorded deaths (95% CI: 38,890 - 40,133 deaths, 4% of deaths, 12 per 100,000) across model runs, the Delta variant caused a median of 273,801 recorded deaths (95% CI: 273,483 - 274,168 deaths, 27% of deaths, 82 per 100,000), and the Omicron variant has so far caused a median of 117,560 recorded deaths (95% CI: 117,252 - 117,835 deaths, 12% of deaths, 35 per 100,000).

**Table 3:**
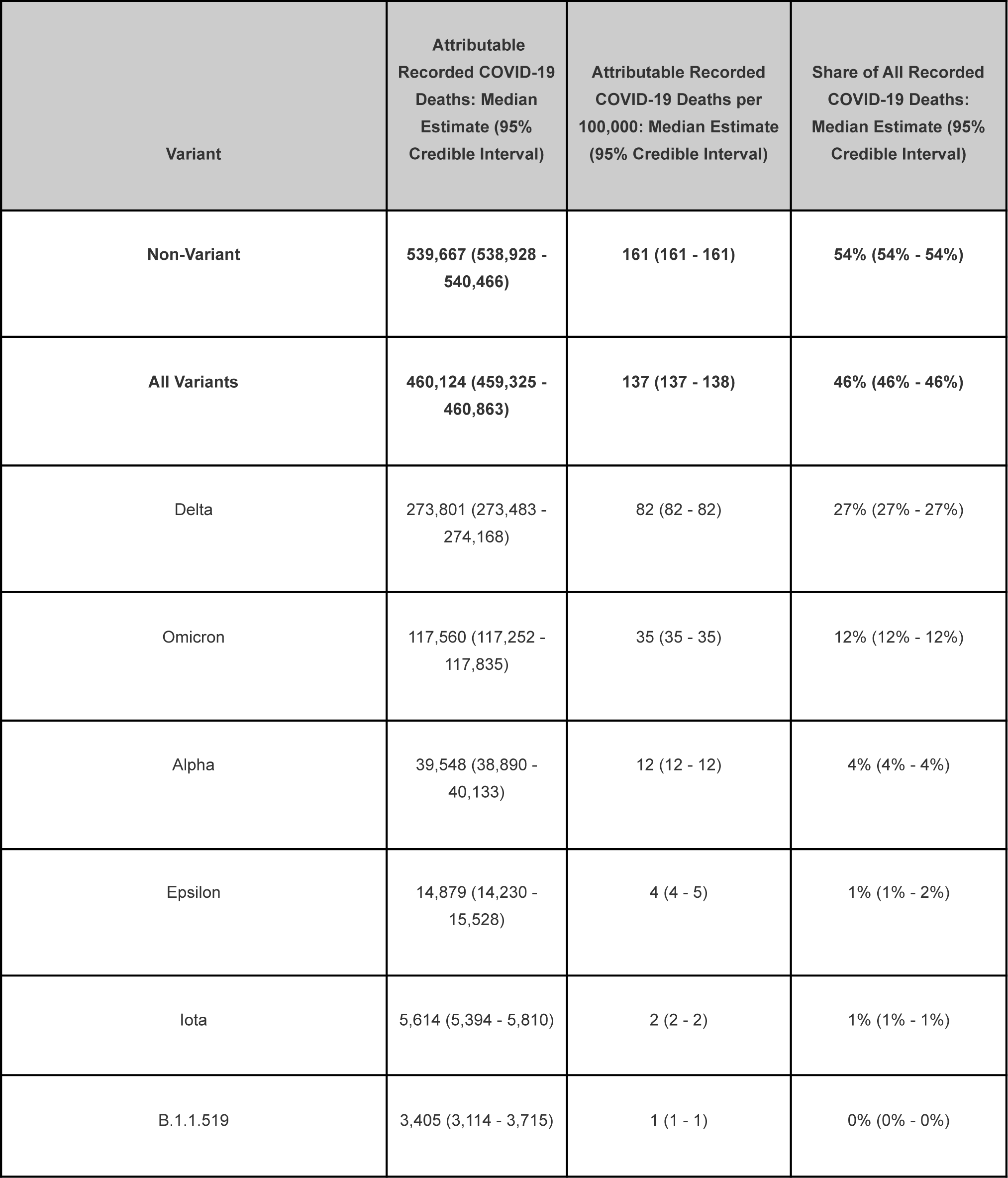
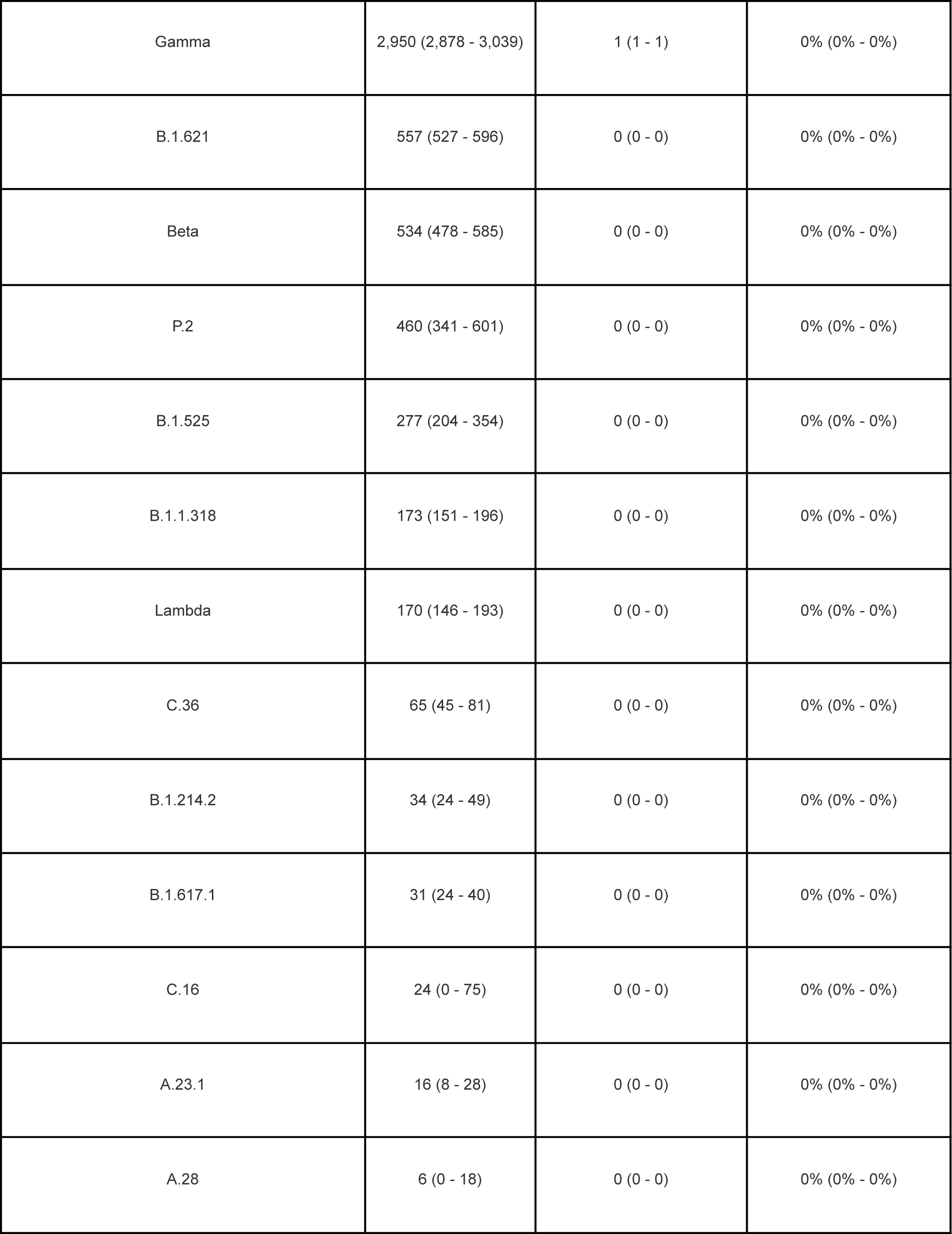
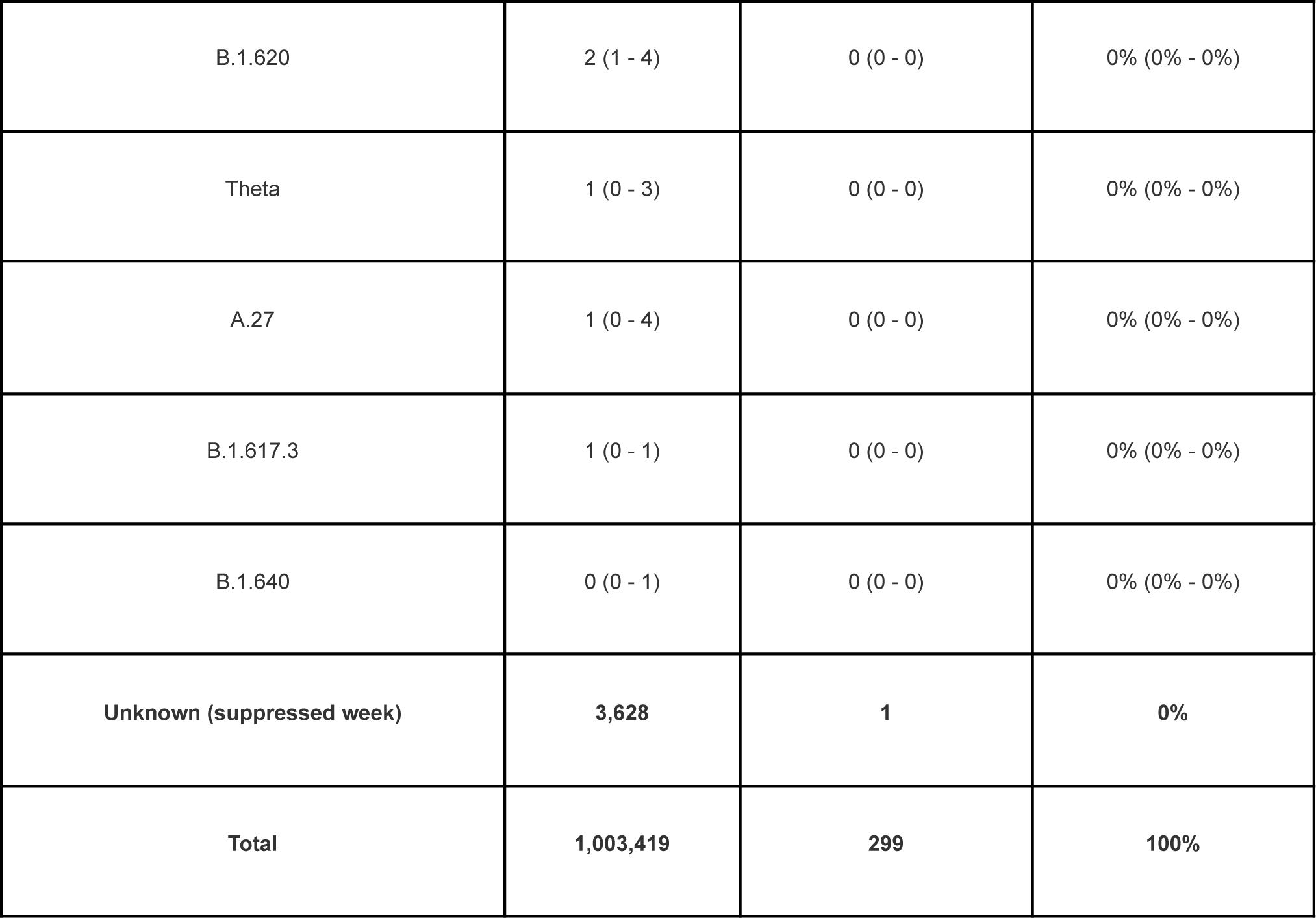
Estimated Number of Recorded COVID-19 Deaths in the U.S. Attributable to Each Variant

In sensitivity analyses where we increased or decreased the assumed severity of Omicron relative to other variants, estimates of recorded deaths caused by Delta ranged from medians of 265,228 deaths (95% CI: 264,898 - 265,600 deaths, 26% of deaths, 79 per 100,000) to 291,946 (95% CI: 291,623 - 292,319 deaths, 29% of deaths, 87 per 100,000) across model runs, while median estimates of recorded deaths caused by Omicron ranged from 99,023 (95% CI: 98,727 - 99,352 deaths, 10% of deaths, 30 per 100,000) to 126,327 (95% CI: 126,022 - 126,582 deaths, 13% of deaths, 38 per 100,000).

Of the COVID-19 deaths recorded by NCHS, 3,628 recorded deaths (0.3%, 1 per 100,000) could not be attributed to a variant because the week of death was not given at the state-level in the dataset. National estimates of the burden of recorded COVID-19 deaths attributable to each variant are given in **Table 3**.

### Regional estimates

The Northeast, South, and Midwest recorded the most COVID-19 deaths per capita (328, 317, and 306 per 100,000 respectively), while the West recorded the fewest (244 per 100,000) [**Figure 1A**]. However, our estimates indicate the Northeast experienced the fewest COVID-19 *variant* deaths per capita (median 111 per 100,000, 95% credible interval: 111 - 112). As a result, variants accounted for 34% of all COVID-19 deaths in the Northeast, compared to nearly half of deaths in each of the other three census regions (Midwest: 46%, South: 50%, West: 49%) [**Tables 4-5**].

**Figure 1:**
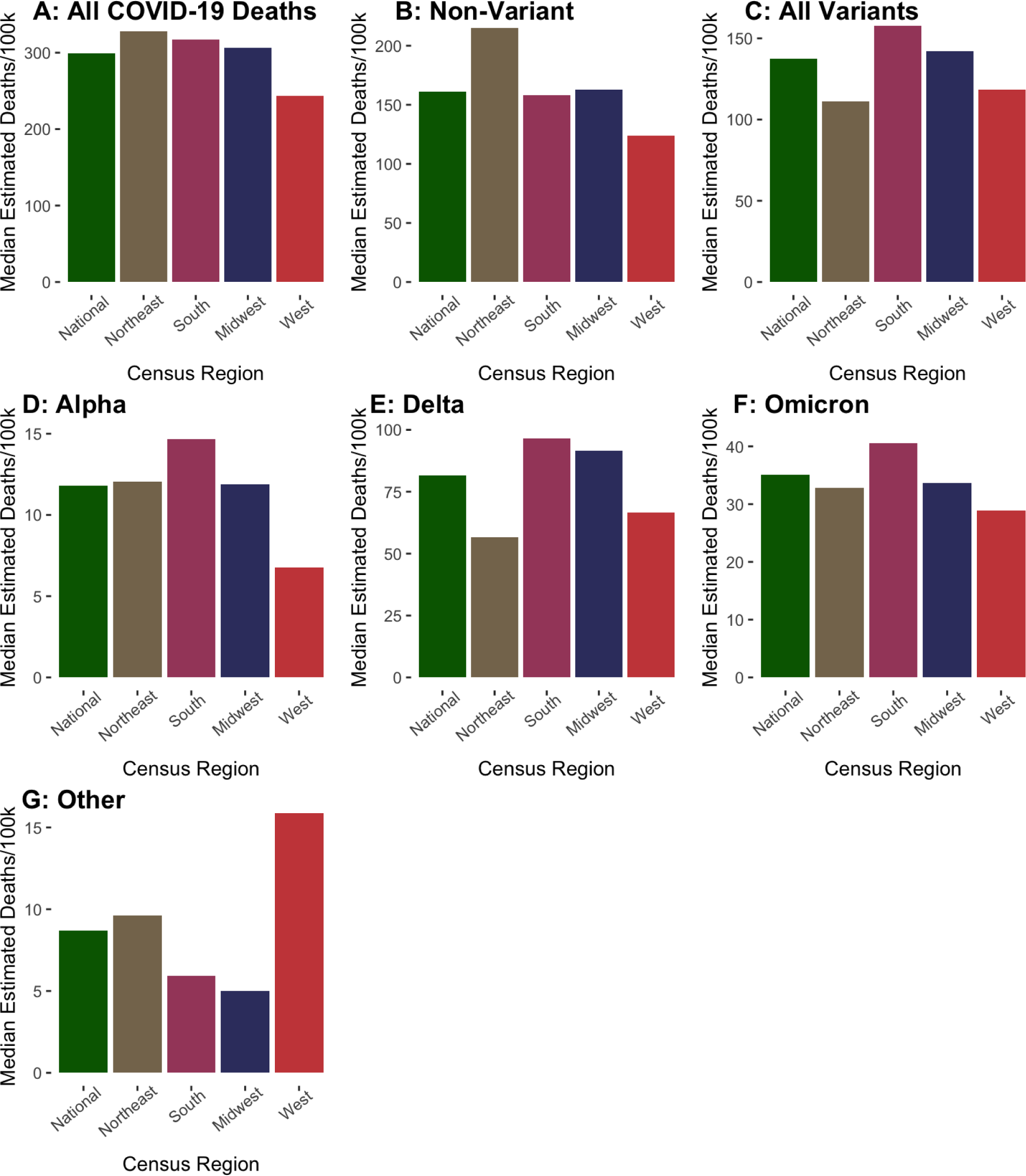
Estimated COVID-19 Deaths per Capita by Variant and U.S. Census Region

#### Non-Variant

The median estimated burden of recorded deaths caused by non-variant SARS-CoV-2 was highest in the Northeast with 215 COVID-19 deaths per 100,000 (123,103 deaths, 66% of all COVID-19 deaths in this region), and lowest in the West with 124 deaths per 100,000 (97,549 deaths, 51% of all COVID-19 deaths in this region) [**Figure 1B, Tables 4-5].** New York City experienced the highest median incidence of deaths from non-variant SARS-CoV-2 (315 deaths per 100,000, or 26,665 deaths overall, 76% of all COVID-19 deaths in NYC) [**Supplementary Tables 1-2**].

**Table 4:** Estimated COVID-19 Deaths per Capita in U.S. by Census Region and Variants, Primary Analysis

| Census Region | Median<br>Estimated<br>Non-Variant<br>COVID-19<br>Deaths per<br>100,000<br>(95% CI, %<br>of Total) | Median<br>Estimated Delta<br>COVID-19<br>Deaths per<br>100,000 (95% CI,<br>% of Total) | Median<br>Estimated<br>Omicron<br>COVID-19<br>Deaths per<br>100,000 (95% CI,<br>% of Total) | Median<br>Estimated Alpha<br>COVID-19 Deaths<br>per 100,000 (95%<br>CI, % of Total) | Median Estimated<br>COVID-19 Deaths<br>per 100,000 from<br>Other Variants<br>(95% CI, % of<br>Total) | Median<br>Estimated<br>COVID-19<br>Deaths per<br>100,000 from All<br>Variants (95% CI,<br>% of Total) | Recorded<br>COVID-19 Deaths<br>per 100,000:<br>Unknown Variant<br>(% of Total) | Total Recorded<br>COVID-19<br>Deaths per<br>100,000 |
| --- | --- | --- | --- | --- | --- | --- | --- | --- |
| National | 161 (161 -<br>161, 54%) | 82 (82 - 82, 27%) | 35 (35 - 35, 12%) | 12 (12 - 12, 4%) | 9 (8 - 9, 3%) | 137 (137 - 138,<br>46%) | 1 (0%) | 299 |
| Northeast (Connecticut,<br>Maine, Massachusetts, New<br>Hampshire, New York,<br>Pennsylvania, Rhode Island,<br>and Vermont) | 215 (215 -<br>216, 66%) | 57 (56 - 57, 17%) | 33 (33 - 33, 10%) | 12 (12 - 12, 4%) | 10 (9 - 10, 3%) | 111 (111 - 112,<br>34%) | 2 (1%) | 328 |
| South (Alabama, Arkansas, Delaware, District of Columbia, Florida, Georgia, Kentucky, Louisiana, Maryland, Mississippi, North Carolina, Oklahoma, Puerto Rico, South Carolina, Tennessee, Texas, Virginia, and West Virginia) | 158 (158 - 159, 50%) | 97 (96 - 97, 30%) | 41 (40 - 41, 13%) | 15 (14 - 15, 5%) | 6 (6 - 6, 2%) | 158 (157 - 158, 50%) | 1 (0%) | 317 |
| Midwest (Illinois, Indiana, Iowa, Kansas, Michigan, Minnesota, Missouri, Nebraska, North Dakota, Ohio, South Dakota, and Wisconsin) | 163 (163 - 163, 53%) | 92 (91 - 92, 30%) | 34 (33 - 34, 11%) | 12 (12 - 12, 4%) | 5 (5 - 5, 2%) | 142 (142 - 143, 46%) | 1 (0%) | 306 |
| West (Alaska, Arizona, California, Colorado, Hawaii, Idaho, Montana, Nevada, New Mexico, Oregon, Utah, Washington, and Wyoming) | 124 (123 - 125, 51%) | 67 (66 - 67, 27%) | 29 (29 - 29, 12%) | 7 (6 - 7, 3%) | 16 (15 - 16, 7%) | 118 (118 - 119, 49%) | 1 (1%) | 244 |

**Table 5:** Estimated COVID-19 Deaths in U.S. by Census Region and Variants, Primary Analysis

| Census Region | Median<br>Estimated<br>Non-Variant<br>COVID-19<br>Deaths (95% CI,<br>% of Total) | Median<br>Estimated Delta<br>COVID-19<br>Deaths (95% CI,<br>% of Total) | Median<br>Estimated<br>Omicron<br>COVID-19<br>Deaths (95% CI,<br>% of Total) | Median Estimated<br>Alpha COVID-19<br>Deaths (95% CI, %<br>of Total) | Median<br>Estimated<br>COVID-19<br>Deaths from<br>Other Variants<br>(95% CI, % of<br>Total) | Median<br>Estimated<br>COVID-19<br>Deaths from<br>All Variants<br>(95% CI, % of<br>Total) | Recorded COVID-19<br>Deaths: Unknown<br>Variant (% of Total) | Total Recorded<br>COVID-19<br>Deaths |
| --- | --- | --- | --- | --- | --- | --- | --- | --- |
| National | 539,667 (538,928 - 540,466, 54%) | 273,801 (273,483 - 274,168, 27%) | 117,560 (117,252 - 117,835, 12%) | 39,548 (38,890 - 40,133, 4%) | 29,208 (28,460 - 29,926, 3%) | 460,124 (459,325 - 460,863, 46%) | 3,628 (0%) | 1,003,419 |
| Northeast (Connecticut, Maine, Massachusetts, New Hampshire, New York, Pennsylvania, Rhode Island, and Vermont) | 123,103 (122,827 - 123,377, 66%) | 32,414 (32,250 - 32,565, 17%) | 18,743 (18,585 - 18,898, 10%) | 6,895 (6,659 - 7,085, 4%) | 5,492 (5,272 - 5,765, 3%) | 63,542 (63,268 - 63,818, 34%) | 991 (1%) | 187,636 |
| South (Alabama, Arkansas, Delaware, District of Columbia, Florida, Georgia, Kentucky, Louisiana, Maryland, Mississippi, North Carolina, Oklahoma, Puerto Rico, South Carolina, Tennessee, Texas, Virginia, and West Virginia) | 206,752 (206,211 - 207,325, 50%) | 125,925 (125,671 - 126,154, 30%) | 52,912 (52,711 - 53,124, 13%) | 19,143 (18,715 - 19,521, 5%) | 7,754 (7,310 - 8,162, 2%) | 205,729 (205,156 - 206,270, 50%) | 828 (0%) | 413,309 |
| Midwest (Illinois, Indiana, Iowa, Kansas, Michigan, Minnesota, Missouri, Nebraska, North Dakota, Ohio, South Dakota, and Wisconsin) | 112,246 (111,992 - 112,480, 53%) | 63,049 (62,910 - 63,222, 30%) | 23,174 (23,015 - 23,324, 11%) | 8,176 (8,031 - 8,316, 4%) | 3,458 (3,245 - 3,678, 2%) | 97,856 (97,622 - 98,110, 46%) | 671 (0%) | 210,773 |
| West (Alaska, Arizona, California, Colorado, Hawaii, Idaho, Montana, Nevada, New Mexico, Oregon, Utah, Washington, and Wyoming) | 97,549 (97,134 - 98,042, 51%) | 52,409 (52,222 - 52,573, 27%) | 22,738 (22,628 - 22,831, 12%) | 5,339 (4,969 - 5,674, 3%) | 12,502 (12,002 - 12,971, 7%) | 93,014 (92,521 - 93,429, 49%) | 1,138 (1%) | 191,701 |

#### Alpha

After its emergence in the winter of 2020/21, the median estimated burden of deaths caused by the Alpha variant was highest in the South with 15 recorded COVID-19 deaths per 100,000 (19,143 deaths, 5% of all COVID-19 deaths in this region) and lowest in the West with 7 recorded COVID-19 deaths per 100,000 (5,339 deaths overall, 3% of all COVID-19 deaths in this region) [**Figure 1D, Tables 4-5**]. Michigan experienced the highest median incidence of Alpha deaths across all jurisdictions (31 deaths per 100,000, or 3,095 deaths overall, 10% of all COVID-19 deaths in Michigan) [**Supplementary Tables 1-2**].

#### Delta

The median estimated burden of deaths caused by the Delta variant was highest in the South with 97 recorded COVID-19 deaths per 100,000 (125,925 deaths, 30% of all COVID-19 deaths in this region), followed by the Midwest with 92 recorded COVID-19 deaths per 100,000 (63,049 deaths, 30% of all COVID-19 deaths in this region). The Northeast experienced the fewest deaths per capita with 57 recorded COVID-19 deaths per 100,000 (32,414 deaths, 17% of all COVID-19 deaths in this region [**Figure 1E, Tables 4-5**]. West Virginia experienced the highest median incidence of Delta deaths (154 per 100,000, or 2,740 deaths overall, 40% of all COVID-19 deaths in West Virginia) across all jurisdictions [**Supplementary Tables 1-2**].

#### Omicron

So far, the median estimated burden of deaths caused by the Omicron variant has been highest in the South with 41 recorded COVID-19 deaths per 100,000 (52,912 deaths, 13% of all COVID-19 deaths in this region) and lowest in the West with 29 recorded COVID-19 deaths per 100,000 (22,738 deaths, 12% of all COVID-19 deaths in this region, respectively) [**Figure 1F, Tables 4-5**]. Mississippi has experienced the highest incidence of Omicron deaths to date (60 per 100,000, or 1,773 deaths overall, 13% of all COVID-19 deaths in Mississippi) across all jurisdictions [**Supplementary Tables 1-2**].

The specific number and share of Omicron and Delta COVID-19 deaths varied in sensitivity analyses where we assumed Omicron was more or less severe (compared to our primary analysis), but the overall regional patterns described above persisted [**Supplementary Tables 3-4**].

#### Other

The median estimated incidence of deaths from other variants (not Alpha, Delta, or Omicron) was higher in the West (16 COVID-19 deaths per 100,000) and Northeast (10 COVID-19 deaths per 100,000) than the Midwest and the South (5 and 6 recorded COVID-19 deaths per 100,000, respectively) [**Figure 1G, Tables 4-5**]. In addition to Alpha, the Epsilon and Iota variants accounted for a significant share of cases in the West and Northeast prior to the emergence of Delta.

## Discussion

Sustained global transmission of SARS-CoV-2 has fueled the emergence of variants that have caused ∼460,000 deaths in the U.S. Alpha, Delta and Omicron—which were first detected in Europe, Asia, and Africa—have led to ∼430,000 deaths so far. The other variants, including those that were first detected in the U.S., accounted for an additional ∼30,000 deaths.

More than 40 percent of national COVID-19 deaths were caused by SARS-CoV-2 variants first detected outside the United States. The phrase “no one is safe until everyone is safe” has been repeatedly invoked as a clarion call during the pandemic. Our analysis provides support for this claim by documenting the significant impact of SARS-CoV-2 variants, which have emerged in the context of uncontrolled transmission both locally and around the world, on U.S. mortality.

Our analysis also suggests the potential benefit of rapidly implementing strategies to reduce the impact of novel variants after the onset of a viral epidemic. We estimate that Delta and Omicron together have caused over 390,000 deaths in the U.S., despite the fact both variants were designated as variants of concern after multiple COVID-19 vaccines had received emergency authorization. Although causality cannot be established definitively, inequalities in global COVID-19 vaccine production and distribution, along with uncontrolled spread of the virus, may have contributed to viral evolution and circulation. For example, Omicron was detected in November 2021 by scientists in Botswana and South Africa, where less than a quarter of the population was fully vaccinated.^32^

Robust global vaccination and treatment campaigns may continue to play an important role in reducing viral replication. A leading hypothesis holds that variants of concern have emerged from immunocompromised individuals with persistent infections that provide more opportunity for viral selection and evolution.^33–35^ While vaccine effectiveness is reduced against Delta and especially Omicron, booster doses provide modest, albeit waning, protection against infection and appear to reduce transmission risk by lowering infectious viral load.^36–39^ When breakthrough cases do occur, vaccinated persons also clear infections faster than unvaccinated persons.^40^ Second-generation vaccines in development could yield significant benefit if they offer greater protection against transmission. In addition, monoclonal antibodies have received emergency authorization for preexposure prophylaxis, and emerging evidence suggests oral antiviral treatments lower SARS-CoV-2 viral load, and clear infections faster.^41–43^

Finally, our analysis reveals the impact of the variants across the country. All regions faced a substantial burden of deaths from the variants. The Northeast recorded the lowest, but still substantial, number of variant deaths, both overall (63,542) and per-capita (111 per 100,000). Some regions were disproportionately impacted, likely reflecting differences in vaccination coverage, prior immunity^44^, use of non-pharmaceutical interventions, demographics and social vulnerability. For example, the South recorded the most variant deaths per capita (median estimate 158 per 100,000) and overall (205,729). It also experienced the highest deaths per capita in each of the Alpha, Delta, and Omicron waves (15, 97 and 41 per 100,000 respectively). Of the 18 states and territories included in the region, 10 had fully vaccinated less than 60% of their population as of early May 2022, according to the CDC.^45^

Our analysis is limited by the quality of public data. NCHS data are considered provisional because of the lag between the date of death and the processing of death certificates by the CDC. Nonetheless, a potential lag would lead to underestimates of deaths caused by variants (due to undercounting of deaths in more recent weeks when Omicron is dominant). In general, reported deaths represent an undercount of total COVID-19 deaths, given limitations in testing and data.^46^ In addition, there is considerable uncertainty associated with some state variant proportions due to insufficient sequencing. Our model also uses variant proportion figures based on clinical testing which may lag behind actual variant circulation. For example, wastewater surveillance suggests that Omicron was circulating in the country before the first case was reported.^47^ This would also lead to underestimates of deaths caused by variants. Finally, we do not model disease transmission dynamics so our estimates should not be interpreted as the number of deaths that would have been prevented if novel variants did not emerge. Even in the absence of new variants, the non-variant SARS-CoV-2 would have continued to spread, resulting in additional deaths.

SARS-CoV-2 variants detected around the world have imposed a significant mortality burden in the U.S. In addition to national public health strategies, greater efforts are needed to reduce the risk of new variants emerging globally. However, U.S. government officials have warned that they are running out of COVID-19 funds.^48^ Congressional lawmakers recently excluded $5 billion in funding for global treatment and vaccination campaigns from a new spending proposal, and there has been limited investment in next-generation vaccines that may offer greater protection against transmission.^49, 50^ Our analysis shows the significant impact of SARS-CoV-2 variants on U.S. public health. In doing so, it highlights the potential risk posed to Americans by new variants–a risk that is only amplified with low rates of global COVID-19 vaccination, and low availability of COVID-19 diagnostics, treatments, and prophylaxis.

## Data Availability

Datasets and R code needed to replicate this analysis are available online at https://github.com/joewalker127/US_Deaths_COVID_Variants

## Acknowledgements

The authors are grateful to Zena Ahmed for research assistance, to Dr. Daniel Weinberger for providing constructive feedback on the analysis, and to Drs. Prabasaj Paul and Molly Steele for assistance in identifying relevant data sources.

## Author Contributions

Concept and design: JW, ZR Statistical analysis: JW

Interpretation of data and findings: JW, NG, GG, VP, ZR

Drafting and revision of the manuscript: JW, NG, GG, VP, ZR

---

**Supplementary Table 1:** Estimated COVID-19 Deaths per Capita in the U.S. by Jurisdiction and Variant, Primary Analysis

| Jurisdiction | Median<br>Estimated<br>Non-Variant<br>COVID-19<br>Deaths per<br>100,000 (95% CI,<br>% of Total) | Median<br>Estimated Delta<br>COVID-19<br>Deaths per<br>100,000 (95% CI,<br>% of Total) | Median<br>Estimated<br>Omicron<br>COVID-19<br>Deaths per<br>100,000 (95% CI,<br>% of Total) | Median<br>Estimated Alpha<br>COVID-19<br>Deaths per<br>100,000 (95% CI,<br>% of Total) | Median<br>Estimated<br>COVID-19<br>Deaths per<br>100,000 from<br>Other Variants<br>(95% CI, % of<br>Total) | Median<br>Estimated<br>COVID-19<br>Deaths per<br>100,000 from All<br>Variants (95%<br>CI, % of Total) | Recorded<br>COVID-19 Deaths<br>per 100,000:<br>Unknown Variant<br>(% of Total) | Total Recorded<br>COVID-19<br>Deaths per<br>100,000 |
| --- | --- | --- | --- | --- | --- | --- | --- | --- |
| Alabama | 206 (205 - 207,<br>54%) | 112 (111 - 113,<br>29%) | 50 (49 - 51, 13%) | 11 (10 - 12, 3%) | 4 (3 - 5, 1%) | 177 (176 - 178,<br>46%) | 0 (0%) | 384 |
| Alaska | 27 (26 - 27, 15%) | 97 (96 - 98, 56%) | 16 (16 - 18, 9%) | 1 (1 - 1, 1%) | 3 (3 - 3, 2%) | 118 (118 - 118,<br>68%) | 30 (17%) | 174 |
| Arizona | 201 (200 - 203,<br>54%) | 112 (111 - 112,<br>30%) | 40 (39 - 40, 11%) | 6 (6 - 7, 2%) | 16 (15 - 18, 4%) | 174 (173 - 175,<br>46%) | 0 (0%) | 375 |
| Arkansas | 194 (193 - 195,<br>53%) | 115 (114 - 117,<br>31%) | 48 (47 - 49, 13%) | 8 (7 - 9, 2%) | 3 (2 - 5, 1%) | 174 (173 - 175,<br>47%) | 1 (0%) | 369 |
| California | 137 (136 - 138,<br>57%) | 46 (46 - 47, 19%) | 28 (28 - 28, 12%) | 7 (6 - 8, 3%) | 23 (22 - 24, 10%) | 105 (104 - 106,<br>43%) | 0 (0%) | 242 |
| Colorado | 107 (105 - 108,<br>47%) | 79 (79 - 80, 35%) | 27 (27 - 28, 12%) | 9 (9 - 9, 4%) | 6 (5 - 7, 2%) | 121 (120 - 123,<br>53%) | 0 (0%) | 228 |
| Connecticut | 216 (215 - 218,<br>71%) | 41 (40 - 41, 13%) | 29 (28 - 30, 9%) | 6 (6 - 7, 2%) | 9 (7 - 10, 3%) | 85 (83 - 86, 28%) | 3 (1%) | 304 |
| Delaware | 153 (152 - 155,<br>53%) | 70 (69 - 72, 24%) | 38 (36 - 39, 13%) | 6 (5 - 7, 2%) | 5 (3 - 6, 2%) | 119 (117 - 120,<br>41%) | 19 (6%) | 291 |
| District of Columbia | 187 (185 - 189, 63%) | 26 (25 - 28, 9%) | 33 (31 - 34, 11%) | 10 (8 - 12, 3%) | 7 (5 - 10, 2%) | 76 (74 - 78, 25%) | 36 (12%) | 298 |
| Florida | 134 (133 - 135, 42%) | 116 (115 - 116, 36%) | 39 (38 - 39, 12%) | 25 (24 - 26, 8%) | 9 (8 - 9, 3%) | 188 (187 - 189, 58%) | 0 (0%) | 322 |
| Georgia | 150 (148 - 152, 50%) | 92 (92 - 93, 31%) | 38 (37 - 38, 13%) | 16 (14 - 18, 5%) | 4 (3 - 5, 1%) | 150 (148 - 152, 50%) | 0 (0%) | 300 |
| Hawaii | 21 (21 - 22, 22%) | 38 (38 - 39, 40%) | 15 (15 - 16, 16%) | 0 (0 - 1, 0%) | 1 (0 - 1, 1%) | 55 (55 - 55, 57%) | 20 (20%) | 96 |
| Idaho | 100 (99 - 102, 38%) | 117 (116 - 118, 45%) | 26 (26 - 27, 10%) | 8 (7 - 9, 3%) | 6 (5 - 8, 2%) | 158 (156 - 159, 60%) | 5 (2%) | 262 |
| Illinois | 162 (161 - 162,<br>60%) | 61 (60 - 62, 23%) | 31 (31 - 32, 12%) | 9 (9 - 10, 3%) | 8 (7 - 8, 3%) | 109 (109 - 110,<br>40%) | 0 (0%) | 271 |
| Indiana | 188 (187 - 189,<br>54%) | 109 (108 - 109,<br>31%) | 37 (36 - 38, 11%) | 10 (10 - 11, 3%) | 5 (4 - 5, 1%) | 161 (160 - 161,<br>46%) | 0 (0%) | 349 |
| Iowa | 182 (180 - 185,<br>61%) | 80 (80 - 81, 27%) | 25 (24 - 26, 8%) | 6 (5 - 6, 2%) | 5 (2 - 7, 2%) | 116 (113 - 119,<br>39%) | 1 (0%) | 300 |
| Kansas | 162 (161 - 163,<br>53%) | 97 (96 - 98, 32%) | 34 (33 - 35, 11%) | 5 (4 - 5, 2%) | 5 (4 - 6, 2%) | 141 (140 - 142,<br>46%) | 2 (1%) | 304 |
| Kentucky | 157 (155 - 158,<br>43%) | 137 (135 - 138,<br>37%) | 57 (56 - 59, 16%) | 12 (11 - 13, 3%) | 5 (4 - 7, 1%) | 212 (210 - 213,<br>57%) | 0 (0%) | 369 |
| Louisiana | 196 (195 - 197,<br>57%) | 90 (89 - 91, 26%) | 37 (36 - 38, 11%) | 13 (12 - 14, 4%) | 8 (7 - 9, 2%) | 147 (146 - 148,<br>43%) | 0 (0%) | 344 |
| Maine | 55 (54 - 55, 30%) | 76 (76 - 77, 42%) | 29 (28 - 30, 16%) | 4 (3 - 4, 2%) | 3 (3 - 4, 2%) | 112 (112 - 113,<br>62%) | 14 (8%) | 181 |
| Maryland | 150 (149 - 152,<br>60%) | 49 (48 - 50, 19%) | 32 (31 - 33, 13%) | 15 (14 - 16, 6%) | 5 (5 - 6, 2%) | 101 (100 - 102,<br>40%) | 0 (0%) | 251 |
| Massachusetts | 186 (185 - 186,<br>71%) | 40 (40 - 40, 15%) | 25 (25 - 26, 10%) | 6 (6 - 7, 2%) | 4 (4 - 5, 2%) | 76 (75 - 76, 29%) | 0 (0%) | 262 |
| Michigan | 151 (151 - 152,<br>48%) | 99 (98 - 100,<br>31%) | 32 (31 - 32, 10%) | 31 (30 - 32, 10%) | 5 (4 - 6, 2%) | 167 (166 - 168,<br>52%) | 0 (0%) | 318 |
| Minnesota | 124 (124 - 124,<br>55%) | 65 (65 - 66, 29%) | 22 (21 - 22, 10%) | 10 (10 - 10, 4%) | 3 (3 - 3, 1%) | 100 (100 - 100,<br>45%) | 0 (0%) | 224 |
| Mississippi | 247 (245 - 249,<br>55%) | 120 (119 - 122,<br>27%) | 60 (59 - 62, 13%) | 14 (12 - 15, 3%) | 5 (3 - 7, 1%) | 199 (197 - 202,<br>45%) | 0 (0%) | 447 |
| Missouri | 171 (170 - 172,<br>51%) | 110 (109 - 111,<br>33%) | 38 (38 - 39, 11%) | 9 (8 - 9, 3%) | 5 (4 - 6, 1%) | 162 (161 - 163,<br>49%) | 0 (0%) | 333 |
| Montana | 137 (135 - 138,<br>43%) | 133 (131 - 134,<br>42%) | 30 (29 - 31, 9%) | 3 (3 - 4, 1%) | 4 (3 - 5, 1%) | 170 (169 - 172,<br>53%) | 12 (4%) | 319 |
| Nebraska | 140 (139 - 141,<br>56%) | 73 (72 - 74, 29%) | 26 (25 - 27, 10%) | 5 (5 - 6, 2%) | 3 (2 - 3, 1%) | 107 (106 - 108,<br>43%) | 4 (2%) | 251 |
| Nevada | 162 (158 - 165,<br>46%) | 116 (115 - 117,<br>33%) | 46 (46 - 47, 13%) | 10 (9 - 10, 3%) | 16 (13 - 20, 5%) | 188 (185 - 192,<br>54%) | 1 (0%) | 351 |
| New<br>Hampshire | 85 (84 - 85, 48%) | 56 (55 - 57, 31%) | 20 (19 - 21, 11%) | 2 (1 - 2, 1%) | 1 (1 - 2, 1%) | 79 (78 - 79, 44%) | 14 (8%) | 178 |
| New Jersey | 241 (240 - 242,<br>70%) | 42 (41 - 43, 12%) | 35 (34 - 35, 10%) | 15 (14 - 17, 4%) | 12 (11 - 13, 3%) | 104 (103 - 105,<br>30%) | 0 (0%) | 345 |
| New Mexico | 187 (186 - 188,<br>51%) | 124 (123 - 126,<br>34%) | 39 (38 - 39, 11%) | 7 (7 - 8, 2%) | 7 (6 - 8, 2%) | 177 (176 - 178,<br>48%) | 3 (1%) | 367 |
| New York<br>State<br>(non-NYC) | 202 (201 - 203,<br>64%) | 57 (56 - 58, 18%) | 35 (35 - 36, 11%) | 11 (11 - 12, 4%) | 11 (11 - 12, 4%) | 115 (114 - 116,<br>36%) | 0 (0%) | 318 |
| New York City | 315 (314 - 316,<br>76%) | 34 (33 - 34, 8%) | 32 (31 - 32, 8%) | 16 (15 - 17, 4%) | 17 (16 - 18, 4%) | 99 (97 - 100,<br>24%) | 0 (0%) | 413 |
| North<br>Carolina | 138 (137 - 139,<br>51%) | 83 (82 - 83, 31%) | 34 (33 - 34, 12%) | 10 (9 - 11, 4%) | 6 (5 - 6, 2%) | 132 (131 - 133,<br>49%) | 0 (0%) | 270 |
| North Dakota | 199 (198 - 200,<br>56%) | 93 (91 - 95, 26%) | 28 (27 - 31, 8%) | 1 (1 - 1, 0%) | 4 (3 - 5, 1%) | 127 (126 - 128,<br>36%) | 31 (9%) | 357 |
| Ohio | 182 (181 - 183,<br>49%) | 123 (122 - 124,<br>33%) | 47 (46 - 48, 13%) | 11 (11 - 12, 3%) | 5 (4 - 6, 1%) | 187 (185 - 188,<br>51%) | 0 (0%) | 369 |
| Oklahoma | 208 (206 - 210,<br>52%) | 125 (123 - 127,<br>31%) | 56 (55 - 58, 14%) | 8 (7 - 9, 2%) | 3 (2 - 5, 1%) | 193 (191 - 194,<br>48%) | 1 (0%) | 402 |
| Oregon | 53 (52 - 53, 31%) | 74 (73 - 74, 44%) | 27 (27 - 28, 16%) | 7 (6 - 7, 4%) | 8 (7 - 9, 5%) | 116 (115 - 117, 69%) | 1 (0%) | 170 |
| Pennsylvania | 199 (198 - 200, 56%) | 96 (95 - 97, 27%) | 38 (38 - 39, 11%) | 15 (15 - 16, 4%) | 7 (7 - 8, 2%) | 157 (156 - 158, 44%) | 0 (0%) | 356 |
| Puerto Rico | 61 (60 - 62, 48%) | 25 (24 - 26, 20%) | 24 (23 - 24, 19%) | 10 (9 - 10, 7%) | 4 (3 - 6, 3%) | 63 (62 - 64, 49%) | 4 (3%) | 129 |
| Rhode Island | 228 (227 - 229, 69%) | 47 (46 - 48, 14%) | 26 (24 - 27, 8%) | 3 (3 - 4, 1%) | 6 (5 - 7, 2%) | 82 (81 - 83, 25%) | 19 (6%) | 329 |
| South Carolina | 178 (177 - 179, 52%) | 105 (105 - 106, 31%) | 45 (44 - 45, 13%) | 10 (10 - 11, 3%) | 6 (5 - 8, 2%) | 167 (166 - 168, 48%) | 0 (0%) | 345 |
| South Dakota | 214 (213 - 215,<br>62%) | 77 (76 - 78, 22%) | 28 (27 - 29, 8%) | 4 (3 - 4, 1%) | 1 (1 - 2, 0%) | 109 (109 - 110,<br>32%) | 20 (6%) | 344 |
| Tennessee | 174 (172 - 175,<br>46%) | 126 (125 - 127,<br>34%) | 55 (54 - 56, 15%) | 17 (17 - 18, 5%) | 4 (3 - 5, 1%) | 202 (201 - 204,<br>54%) | 0 (0%) | 376 |
| Texas | 171 (170 - 173,<br>53%) | 92 (91 - 92, 28%) | 38 (38 - 38, 12%) | 14 (13 - 15, 4%) | 7 (6 - 8, 2%) | 151 (149 - 152,<br>47%) | 0 (0%) | 322 |
| Utah | 68 (68 - 69, 44%) | 59 (59 - 60, 39%) | 17 (17 - 17, 11%) | 3 (3 - 3, 2%) | 4 (3 - 5, 3%) | 83 (83 - 84, 54%) | 2 (1%) | 154 |
| Vermont | 16 (16 - 16, 17%) | 27 (26 - 28, 27%) | 16 (15 - 17, 16%) | 0 (0 - 0, 0%) | 0 (0 - 0, 0%) | 42 (42 - 42, 44%) | 38 (39%) | 97 |
| Virginia | 117 (116 - 117,<br>52%) | 61 (60 - 62, 27%) | 33 (32 - 33, 15%) | 9 (9 - 9, 4%) | 4 (4 - 5, 2%) | 107 (107 - 108,<br>48%) | 0 (0%) | 224 |
| Washington | 63 (62 - 64, 41%) | 57 (56 - 57, 36%) | 24 (24 - 24, 15%) | 6 (5 - 6, 4%) | 7 (6 - 7, 4%) | 93 (92 - 94, 59%) | 0 (0%) | 156 |
| West Virginia | 150 (149 - 151,<br>39%) | 154 (152 - 155,<br>40%) | 58 (57 - 60, 15%) | 13 (12 - 14, 3%) | 4 (3 - 5, 1%) | 229 (228 - 230,<br>60%) | 6 (1%) | 384 |
| Wisconsin | 134 (133 - 134,<br>54%) | 80 (79 - 80, 32%) | 27 (26 - 27, 11%) | 5 (5 - 5, 2%) | 4 (3 - 4, 1%) | 115 (114 - 116,<br>46%) | 0 (0%) | 249 |
| Wyoming | 91 (90 - 92, 33%) | 130 (128 - 132,<br>47%) | 24 (23 - 26, 9%) | 0 (0 - 1, 0%) | 3 (2 - 4, 1%) | 157 (156 - 158,<br>56%) | 31 (11%) | 280 |

**Supplementary Table 2:** Estimated COVID-19 Deaths in the U.S. by Jurisdiction and Variant, Primary Analysis

| Jurisdiction | Median Estimated<br>Non-Variant<br>COVID-19 Deaths<br>(95% CI, % of<br>Total) | Median<br>Estimated<br>Delta<br>COVID-19<br>Deaths (95%<br>CI, % of Total) | Median<br>Estimated<br>Omicron<br>COVID-19 Deaths<br>(95% CI, % of<br>Total) | Median Estimated<br>Alpha COVID-19<br>Deaths (95% CI,<br>% of Total) | Median Estimated<br>COVID-19 Deaths<br>from Other<br>Variants (95% CI,<br>% of Total) | Median Estimated<br>COVID-19 Deaths<br>from All Variants<br>(95% CI, % of<br>Total) | Recorded<br>COVID-19<br>Deaths:<br>Unknown Variant<br>(% of Total) | Total Recorded<br>COVID-19<br>Deaths |
| --- | --- | --- | --- | --- | --- | --- | --- | --- |
| Alabama | 10,402 (10,347 -<br>10,449, 54%) | 5,660 (5,610 -<br>5,715, 29%) | 2,534 (2,492 -<br>2,579, 13%) | 547 (500 - 604,<br>3%) | 181 (140 - 242, 1%) | 8,925 (8,878 -<br>8,980, 46%) | 10 (0%) | 19,337 |
| Alaska | 195 (194 - 196,<br>15%) | 714 (704 - 721,<br>56%) | 120 (114 - 130,<br>9%) | 8 (5 - 11, 1%) | 21 (18 - 24, 2%) | 863 (862 - 864,<br>68%) | 217 (17%) | 1,275 |
| Arizona | 14,659 (14,543 -<br>14,754, 54%) | 8,115 (8,065 -<br>8,157, 30%) | 2,899 (2,861 -<br>2,946, 11%) | 460 (430 - 487,<br>2%) | 1,176 (1,079 -<br>1,296, 4%) | 12,647 (12,552 -<br>12,763, 46%) | 15 (0%) | 27,321 |
| Arkansas | 5,880 (5,842 - 5,909, 53%) | 3,488 (3,436 - 3,526, 31%) | 1,440 (1,408 - 1,483, 13%) | 228 (202 - 258, 2%) | 100 (73 - 142, 1%) | 5,256 (5,227 - 5,294, 47%) | 23 (0%) | 11,159 |
| California | 53,817 (53,410 - 54,230, 57%) | 18,219 (18,075 - 18,373, 19%) | 11,085 (11,025 - 11,142, 12%) | 2,866 (2,503 - 3,197, 3%) | 9,127 (8,747 - 9,594, 10%) | 41,300 (40,887 - 41,707, 43%) | 13 (0%) | 95,130 |
| Colorado | 6,199 (6,128 - 6,261, 47%) | 4,598 (4,564 - 4,632, 35%) | 1,591 (1,560 - 1,625, 12%) | 533 (519 - 549, 4%) | 329 (271 - 400, 2%) | 7,052 (6,990 - 7,123, 53%) | 14 (0%) | 13,265 |
| Connecticut | 7,793 (7,736 - 7,849, 71%) | 1,466 (1,433 - 1,491, 13%) | 1,036 (1,011 - 1,065, 9%) | 234 (220 - 250, 2%) | 315 (262 - 378, 3%) | 3,052 (2,996 - 3,109, 28%) | 103 (1%) | 10,948 |
| Delaware | 1,539 (1,522 - 1,554, 53%) | 705 (690 - 720, 24%) | 379 (365 - 394, 13%) | 59 (50 - 69, 2%) | 46 (31 - 61, 2%) | 1,190 (1,175 - 1,207, 41%) | 189 (6%) | 2,918 |
| District of Columbia | 1,252 (1,239 - 1,268, 63%) | 177 (165 - 190, 9%) | 219 (206 - 231, 11%) | 68 (55 - 80, 3%) | 45 (31 - 64, 2%) | 509 (493 - 522, 25%) | 238 (12%) | 1,999 |
| Florida | 29,179 (28,976 - 29,399, 42%) | 25,174 (25,093 - 25,251, 36%) | 8,428 (8,371 - 8,482, 12%) | 5,422 (5,230 - 5,626, 8%) | 1,919 (1,837 - 2,030, 3%) | 40,941 (40,721 - 41,144, 58%) | 4 (0%) | 70,124 |
| Georgia | 16,226 (16,016 - 16,401, 50%) | 9,966 (9,911 - 10,014, 31%) | 4,061 (4,017 - 4,110, 13%) | 1,705 (1,542 - 1,905, 5%) | 455 (365 - 546, 1%) | 16,194 (16,019 - 16,404, 50%) | 15 (0%) | 32,435 |
| Hawaii | 309 (304 - 314, 22%) | 554 (546 - 562, 40%) | 221 (214 - 230, 16%) | 6 (4 - 8, 0%) | 12 (7 - 17, 1%) | 793 (788 - 798, 57%) | 284 (20%) | 1,386 |
| Idaho | 1,899 (1,874 - 1,930, 38%) | 2,227 (2,212 - 2,241, 45%) | 500 (489 - 511, 10%) | 152 (137 - 169, 3%) | 119 (92 - 147, 2%) | 2,998 (2,967 - 3,023, 60%) | 92 (2%) | 4,989 |
| Illinois | 20,499 (20,401 - 20,578, 60%) | 7,732 (7,656 - 7,796, 23%) | 3,952 (3,890 - 4,017, 12%) | 1,189 (1,149 - 1,237, 3%) | 963 (882 - 1,054, 3%) | 13,837 (13,758 - 13,935, 40%) | 1 (0%) | 34,337 |
| Indiana | 12,789 (12,731 - 12,846, 54%) | 7,390 (7,327 - 7,439, 31%) | 2,518 (2,469 - 2,572, 11%) | 704 (660 - 754, 3%) | 312 (273 - 365, 1%) | 10,927 (10,870 - 10,985, 46%) | 18 (0%) | 23,734 |
| Iowa | 5,818 (5,736 - 5,903, 61%) | 2,564 (2,541 - 2,585, 27%) | 798 (781 - 817, 8%) | 186 (171 - 201, 2%) | 157 (71 - 238, 2%) | 3,704 (3,619 - 3,786, 39%) | 42 (0%) | 9,564 |
| Kansas | 4,754 (4,722 - 4,778, 53%) | 2,849 (2,823 - 2,874, 32%) | 1,002 (979 - 1,025, 11%) | 134 (120 - 149, 2%) | 144 (120 - 177, 2%) | 4,128 (4,104 - 4,160, 46%) | 50 (1%) | 8,932 |
| Kentucky | 7,075 (7,005 - 7,142, 43%) | 6,180 (6,108 - 6,245, 37%) | 2,591 (2,533 - 2,656, 16%) | 550 (504 - 599, 3%) | 221 (167 - 299, 1%) | 9,546 (9,479 - 9,616, 57%) | 20 (0%) | 16,641 |
| Louisiana | 9,076 (9,030 - 9,126, 57%) | 4,151 (4,107 - 4,198, 26%) | 1,714 (1,683 - 1,745, 11%) | 605 (565 - 648, 4%) | 348 (303 - 396, 2%) | 6,819 (6,769 - 6,865, 43%) | 18 (0%) | 15,913 |
| Maine | 748 (742 - 756, 30%) | 1,048 (1,037 - 1,059, 42%) | 395 (384 - 406, 16%) | 50 (44 - 57, 2%) | 46 (41 - 52, 2%) | 1,540 (1,532 - 1,546, 62%) | 192 (8%) | 2,480 |
| Maryland | 9,277 (9,192 - 9,346, 60%) | 3,022 (2,975 - 3,075, 19%) | 1,957 (1,905 - 2,009, 13%) | 899 (844 - 986, 6%) | 331 (284 - 369, 2%) | 6,209 (6,140 - 6,294, 40%) | 12 (0%) | 15,498 |
| Massachusetts | 12,966 (12,917 - 13,009, 71%) | 2,788 (2,759 - 2,813, 15%) | 1,767 (1,743 - 1,793, 10%) | 432 (399 - 466, 2%) | 296 (266 - 326, 2%) | 5,281 (5,238 - 5,330, 29%) | 25 (0%) | 18,272 |
| Michigan | 15,227 (15,142 - 15,322, 48%) | 9,961 (9,897 - 10,019, 31%) | 3,191 (3,136 - 3,240, 10%) | 3,095 (3,012 - 3,179, 10%) | 512 (446 - 587, 2%) | 16,761 (16,666 - 16,846, 52%) | 4 (0%) | 31,992 |
| Minnesota | 7,072 (7,053 - 7,089, 55%) | 3,718 (3,694 - 3,741, 29%) | 1,247 (1,225 - 1,271, 10%) | 565 (551 - 577, 4%) | 166 (153 - 180, 1%) | 5,696 (5,679 - 5,715, 45%) | 1 (0%) | 12,769 |
| Mississippi | 7,290 (7,226 - 7,348, 55%) | 3,551 (3,502 - 3,605, 27%) | 1,773 (1,728 - 1,831, 13%) | 404 (360 - 443, 3%) | 156 (95 - 217, 1%) | 5,884 (5,826 - 5,948, 45%) | 10 (0%) | 13,184 |
| Missouri | 10,560 (10,466 - 10,626, 51%) | 6,804 (6,753 - 6,856, 33%) | 2,356 (2,315 - 2,402, 11%) | 528 (491 - 569, 3%) | 285 (217 - 370, 1%) | 9,974 (9,908 - 10,068, 49%) | 10 (0%) | 20,544 |
| Montana | 1,511 (1,495 - 1,523, 43%) | 1,464 (1,449 - 1,482, 42%) | 333 (315 - 347, 9%) | 37 (29 - 45, 1%) | 44 (32 - 60, 1%) | 1,878 (1,866 - 1,894, 53%) | 137 (4%) | 3,526 |
| Nebraska | 2,751 (2,731 - 2,768, 56%) | 1,436 (1,421 - 1,451, 29%) | 512 (497 - 527, 10%) | 102 (94 - 110, 2%) | 50 (33 - 69, 1%) | 2,099 (2,082 - 2,119, 43%) | 87 (2%) | 4,937 |
| Nevada | 5,082 (4,971 - 5,176, 46%) | 3,641 (3,615 - 3,670, 33%) | 1,460 (1,435 - 1,483, 13%) | 305 (276 - 328, 3%) | 513 (403 - 624, 5%) | 5,918 (5,824 - 6,029, 54%) | 23 (0%) | 11,023 |
| New Hampshire | 1,176 (1,167 - 1,184, 48%) | 775 (760 - 789, 31%) | 276 (261 - 289, 11%) | 24 (21 - 31, 1%) | 18 (13 - 26, 1%) | 1,093 (1,085 - 1,102, 44%) | 201 (8%) | 2,470 |
| New Jersey | 22,335 (22,202 - 22,450, 70%) | 3,884 (3,823 - 3,943, 12%) | 3,210 (3,148 - 3,266, 10%) | 1,436 (1,332 - 1,529, 4%) | 1,106 (1,033 - 1,212, 3%) | 9,635 (9,520 - 9,768, 30%) | 3 (0%) | 31,973 |
| New Mexico | 3,958 (3,935 - 3,981, 51%) | 2,634 (2,612 - 2,656, 34%) | 815 (798 - 835, 11%) | 153 (140 - 168, 2%) | 147 (124 - 171, 2%) | 3,751 (3,728 - 3,774, 48%) | 53 (1%) | 7,762 |
| New York State<br>(non-NYC) | 22,990 (22,865 - 23,110, 64%) | 6,484 (6,413 - 6,552, 18%) | 4,012 (3,953 - 4,090, 11%) | 1,305 (1,215 - 1,404, 4%) | 1,300 (1,203 - 1,397, 4%) | 13,102 (12,982 - 13,227, 36%) | 9 (0%) | 36,101 |
| New York City | 26,665 (26,571 - 26,786, 76%) | 2,849 (2,795 - 2,904, 8%) | 2,686 (2,637 - 2,740, 8%) | 1,386 (1,301 - 1,470, 4%) | 1,425 (1,339 - 1,517, 4%) | 8,346 (8,225 - 8,440, 24%) | 2 (0%) | 35,013 |
| North Carolina | 14,590 (14,484 - 14,683, 51%) | 8,722 (8,676 - 8,765, 31%) | 3,540 (3,502 - 3,581, 12%) | 1,074 (996 - 1,169, 4%) | 582 (525 - 666, 2%) | 13,924 (13,831 - 14,030, 49%) | 4 (0%) | 28,518 |
| North Dakota | 1,542 (1,533 - 1,550, 56%) | 724 (704 - 739, 26%) | 221 (207 - 240, 8%) | 8 (5 - 11, 0%) | 33 (26 - 42, 1%) | 986 (978 - 995, 36%) | 237 (9%) | 2,765 |
| Ohio | 21,444 (21,277 - 21,606, 49%) | 14,499 (14,391 - 14,623, 33%) | 5,535 (5,420 - 5,647, 13%) | 1,346 (1,275 - 1,429, 3%) | 607 (473 - 747, 1%) | 21,985 (21,823 - 22,152, 51%) | 11 (0%) | 43,440 |
| Oklahoma | 8,299 (8,231 - 8,365, 52%) | 4,993 (4,921 - 5,059, 31%) | 2,241 (2,184 - 2,299, 14%) | 318 (275 - 373, 2%) | 129 (91 - 185, 1%) | 7,686 (7,620 - 7,754, 48%) | 24 (0%) | 16,009 |
| Oregon | 2,237 (2,201 - 2,269, 31%) | 3,132 (3,103 - 3,156, 44%) | 1,154 (1,132 - 1,177, 16%) | 292 (269 - 311, 4%) | 354 (318 - 395, 5%) | 4,932 (4,900 - 4,968, 69%) | 29 (0%) | 7,198 |
| Pennsylvania | 25,834 (25,713 - 25,929, 56%) | 12,438 (12,334 - 12,527, 27%) | 4,971 (4,889 - 5,070, 11%) | 1,978 (1,913 - 2,069, 4%) | 918 (846 - 1,007, 2%) | 20,312 (20,217 - 20,433, 44%) | 5 (0%) | 46,151 |
| Puerto Rico | 1,998 (1,956 - 2,037, 48%) | 819 (797 - 835, 20%) | 778 (762 - 799, 19%) | 310 (290 - 330, 7%) | 146 (110 - 186, 3%) | 2,052 (2,013 - 2,094, 49%) | 144 (3%) | 4,194 |
| Rhode Island | 2,499 (2,485 - 2,513, 69%) | 518 (505 - 531, 14%) | 280 (266 - 292, 8%) | 37 (33 - 42, 1%) | 64 (50 - 77, 2%) | 898 (884 - 912, 25%) | 204 (6%) | 3,601 |
| South Carolina | 9,236 (9,169 - 9,293, 52%) | 5,475 (5,430 - 5,512, 31%) | 2,323 (2,289 - 2,361, 13%) | 543 (494 - 590, 3%) | 329 (266 - 396, 2%) | 8,665 (8,608 - 8,732, 48%) | 4 (0%) | 17,905 |
| South Dakota | 1,917 (1,911 - 1,924, 62%) | 689 (679 - 701, 22%) | 250 (238 - 261, 8%) | 31 (26 - 37, 1%) | 10 (6 - 13, 0%) | 980 (973 - 986, 32%) | 182 (6%) | 3,079 |
| Tennessee | 12,117 (12,004 - 12,187, 46%) | 8,791 (8,723 - 8,853, 34%) | 3,818 (3,763 - 3,875, 15%) | 1,217 (1,163 - 1,261, 5%) | 278 (215 - 377, 1%) | 14,102 (14,032 - 14,215, 54%) | 3 (0%) | 26,222 |
| Texas | 50,572 (50,151 - 50,960, 53%) | 27,045 (26,966 - 27,122, 28%) | 11,246 (11,178 - 11,320, 12%) | 4,159 (3,918 - 4,395, 4%) | 2,003 (1,690 - 2,369, 2%) | 44,448 (44,060 - 44,869, 47%) | 2 (0%) | 95,022 |
| Utah | 2,278 (2,254 - 2,299, 44%) | 1,975 (1,955 - 1,993, 39%) | 568 (555 - 584, 11%) | 103 (89 - 116, 2%) | 129 (105 - 152, 3%) | 2,775 (2,754 - 2,799, 54%) | 72 (1%) | 5,125 |
| Vermont | 106 (106 - 106, 17%) | 171 (165 - 178, 27%) | 103 (96 - 108, 16%) | 0 (0 - 0, 0%) | 0 (0 - 0, 0%) | 274 (274 - 274, 44%) | 247 (39%) | 627 |
| Virginia | 10,102 (10,055 - 10,149, 52%) | 5,270 (5,216 - 5,326, 27%) | 2,831 (2,777 - 2,881, 15%) | 790 (759 - 820, 4%) | 381 (342 - 425, 2%) | 9,270 (9,223 - 9,317, 48%) | 8 (0%) | 19,380 |
| Washington | 4,899 (4,829 - 4,958, 41%) | 4,386 (4,352 - 4,416, 36%) | 1,850 (1,824 - 1,882, 15%) | 429 (406 - 454, 4%) | 504 (448 - 576, 4%) | 7,173 (7,114 - 7,243, 59%) | 11 (0%) | 12,083 |
| West Virginia | 2,672 (2,654 - 2,691, 39%) | 2,740 (2,709 - 2,770, 40%) | 1,036 (1,011 - 1,068, 15%) | 230 (216 - 244, 3%) | 72 (57 - 87, 1%) | 4,079 (4,060 - 4,097, 60%) | 100 (1%) | 6,851 |
| Wisconsin | 7,877 (7,828 - 7,913, 54%) | 4,694 (4,664 - 4,723, 32%) | 1,584 (1,558 - 1,611, 11%) | 287 (271 - 305, 2%) | 210 (171 - 257, 1%) | 6,775 (6,739 - 6,824, 46%) | 28 (0%) | 14,680 |
| Wyoming | 529 (523 - 535, 33%) | 754 (743 - 763, 47%) | 138 (131 - 150, 9%) | 1 (0 - 4, 0%) | 17 (11 - 23, 1%) | 911 (905 - 917, 56%) | 178 (11%) | 1,618 |

**Supplementary Table 3:**
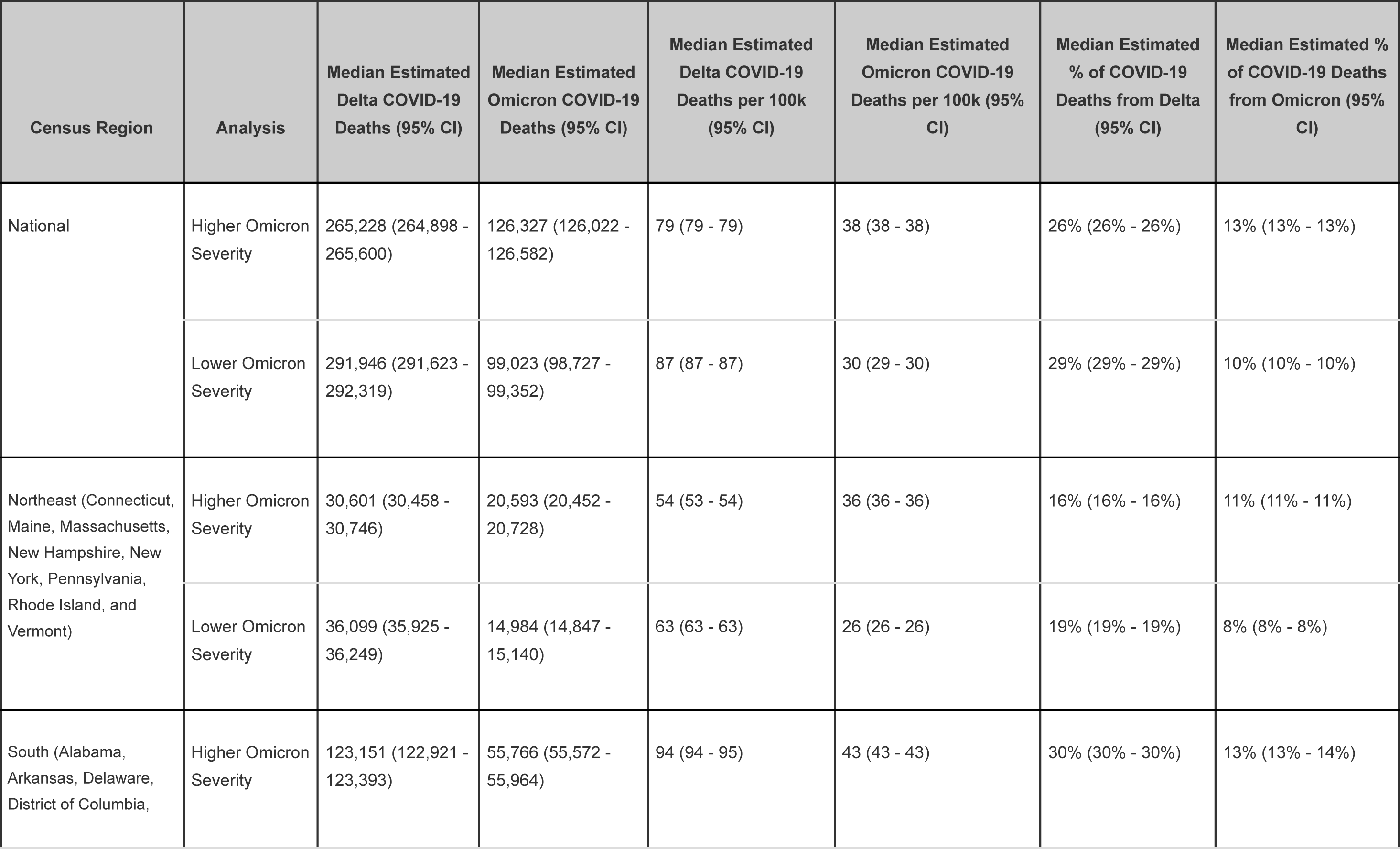
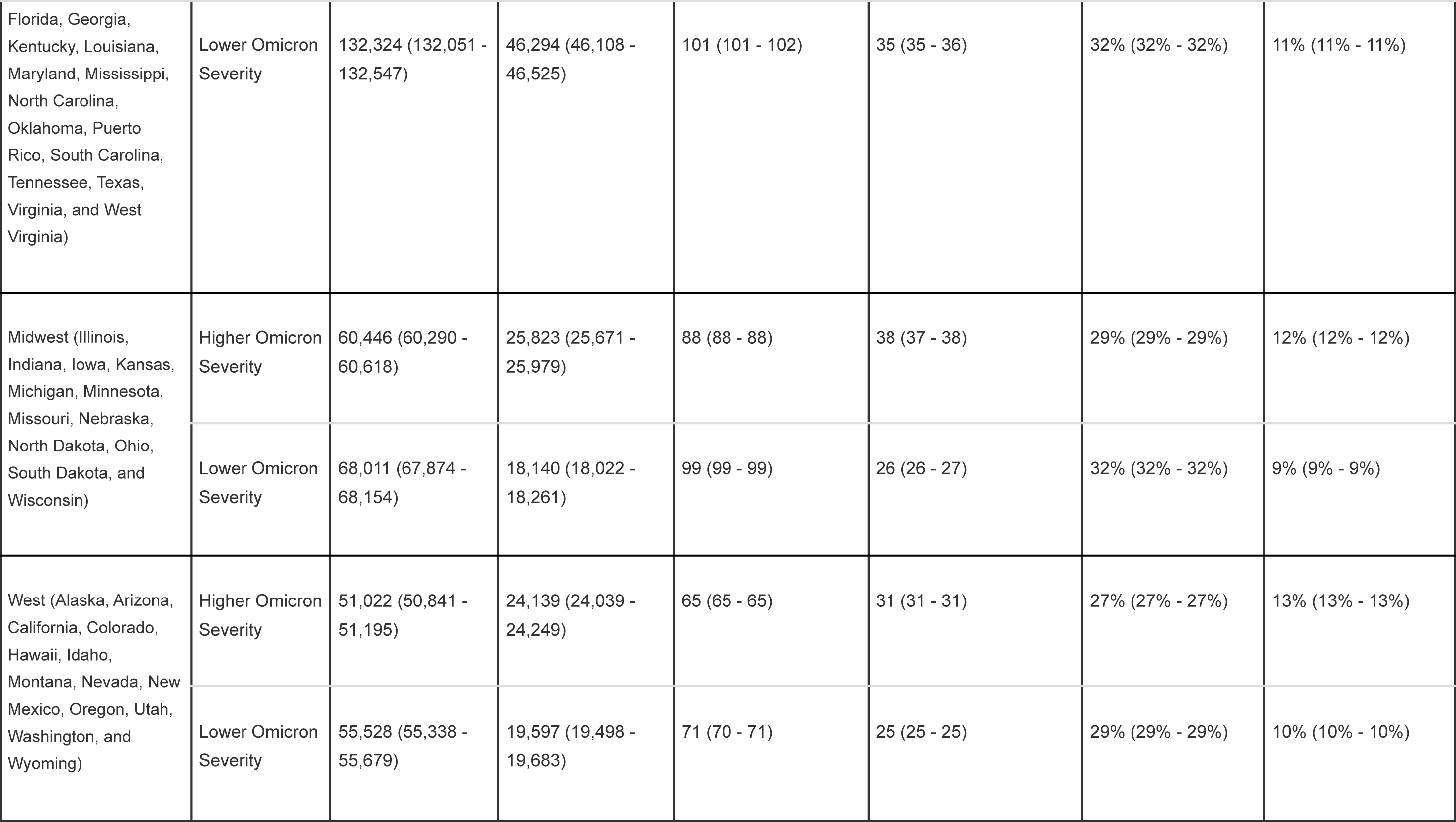
Estimated Omicron and Delta COVID-19 Deaths in U.S. by Census Region, Sensitivity Analysis Assuming Higher or Lower Omicron Severity

**Supplementary Table 4:** Estimated Omicron and Delta COVID-19 Deaths in U.S. by Jurisdiction, Sensitivity Analysis Assuming Higher or Lower Omicron Severity

| Jurisdiction | Analysis | Median Estimated<br>Delta COVID-19<br>Deaths (95% CI) | Median Estimated<br>Omicron COVID-19<br>Deaths (95% CI) | Median Estimated<br>Delta COVID-19<br>Deaths per 100k (95%<br>CI) | Median Estimated<br>Omicron COVID-19<br>Deaths per 100k (95%<br>CI) | Median Estimated<br>% of COVID-19<br>Deaths from Delta<br>(95% CI) | Median Estimated<br>% of COVID-19<br>Deaths from<br>Omicron (95% CI) |
| --- | --- | --- | --- | --- | --- | --- | --- |
| Alabama | Higher<br>Omicron<br>Severity | 5,572 (5,528 -<br>5,624) | 2,624 (2,585 - 2,664) | 111 (110 - 112) | 52 (51 - 53) | 29% (29% - 29%) | 14% (13% - 14%) |
| Alabama | Lower<br>Omicron<br>Severity | 5,893 (5,826 -<br>5,954) | 2,295 (2,242 - 2,357) | 117 (116 - 118) | 46 (44 - 47) | 30% (30% - 31%) | 12% (12% - 12%) |
| Alaska | Higher<br>Omicron<br>Severity | 707 (699 - 715) | 127 (120 - 134) | 96 (95 - 98) | 17 (16 - 18) | 55% (55% - 56%) | 10% (9% - 11%) |
| Alaska | Lower<br>Omicron<br>Severity | 730 (717 - 737) | 104 (98 - 117) | 100 (98 - 101) | 14 (13 - 16) | 57% (56% - 58%) | 8% (8% - 9%) |
| Arizona | Higher<br>Omicron<br>Severity | 7,922 (7,870 -<br>7,965) | 3,094 (3,052 - 3,146) | 109 (108 - 109) | 43 (42 - 43) | 29% (29% - 29%) | 11% (11% - 12%) |
| Arizona | Lower<br>Omicron<br>Severity | 8,483 (8,440 -<br>8,517) | 2,530 (2,499 - 2,562) | 117 (116 - 117) | 35 (34 - 35) | 31% (31% - 31%) | 9% (9% - 9%) |
| Arkansas | Higher<br>Omicron<br>Severity | 3,412 (3,360 -<br>3,453) | 1,516 (1,482 - 1,559) | 113 (111 - 114) | 50 (49 - 52) | 31% (30% - 31%) | 14% (13% - 14%) |
| Arkansas | Lower<br>Omicron<br>Severity | 3,647 (3,604 -<br>3,676) | 1,281 (1,258 - 1,314) | 121 (119 - 121) | 42 (42 - 43) | 33% (32% - 33%) | 11% (11% - 12%) |
| California | Higher<br>Omicron<br>Severity | 17,572 (17,435 -<br>17,726) | 11,739 (11,685 -<br>11,807) | 45 (44 - 45) | 30 (30 - 30) | 18% (18% - 19%) | 12% (12% - 12%) |
| California | Lower<br>Omicron<br>Severity | 19,749 (19,605 -<br>19,905) | 9,545 (9,481 - 9,593) | 50 (50 - 51) | 24 (24 - 24) | 21% (21% - 21%) | 10% (10% - 10%) |
| Colorado | Higher<br>Omicron<br>Severity | 4,481 (4,451 - 4,512) | 1,708 (1,676 - 1,735) | 77 (77 - 78) | 29 (29 - 30) | 34% (34% - 34%) | 13% (13% - 13%) |
| Colorado | Lower<br>Omicron<br>Severity | 4,844 (4,808 - 4,879) | 1,342 (1,313 - 1,376) | 83 (83 - 84) | 23 (23 - 24) | 37% (36% - 37%) | 10% (10% - 10%) |
| Connecticut | Higher<br>Omicron<br>Severity | 1,361 (1,330 - 1,386) | 1,143 (1,117 - 1,172) | 38 (37 - 38) | 32 (31 - 33) | 12% (12% - 13%) | 10% (10% - 11%) |
| Connecticut | Lower<br>Omicron<br>Severity | 1,691 (1,664 - 1,716) | 806 (784 - 834) | 47 (46 - 48) | 22 (22 - 23) | 15% (15% - 16%) | 7% (7% - 8%) |
| Delaware | Higher<br>Omicron<br>Severity | 671 (655 - 684) | 413 (400 - 428) | 67 (65 - 68) | 41 (40 - 43) | 23% (22% - 23%) | 14% (14% - 15%) |
| Delaware | Lower<br>Omicron<br>Severity | 780 (762 - 795) | 303 (290 - 321) | 78 (76 - 79) | 30 (29 - 32) | 27% (26% - 27%) | 10% (10% - 11%) |
| District of Columbia | Higher Omicron Severity | 159 (149 - 170) | 236 (224 - 247) | 24 (22 - 25) | 35 (33 - 37) | 8% (7% - 9%) | 12% (11% - 12%) |
| District of Columbia | Lower Omicron Severity | 222 (209 - 238) | 173 (158 - 187) | 33 (31 - 36) | 26 (24 - 28) | 11% (10% - 12%) | 9% (8% - 9%) |
| Florida | Higher Omicron Severity | 24,814 (24,734 - 24,888) | 8,806 (8,760 - 8,852) | 114 (114 - 114) | 40 (40 - 41) | 35% (35% - 35%) | 13% (12% - 13%) |
| Florida | Lower Omicron Severity | 26,101 (26,007 - 26,203) | 7,449 (7,371 - 7,520) | 120 (119 - 120) | 34 (34 - 35) | 37% (37% - 37%) | 11% (11% - 11%) |
| Georgia | Higher Omicron Severity | 9,754 (9,706 - 9,801) | 4,281 (4,246 - 4,322) | 90 (90 - 91) | 40 (39 - 40) | 30% (30% - 30%) | 13% (13% - 13%) |
| Georgia | Lower Omicron Severity | 10,481 (10,419 - 10,540) | 3,520 (3,465 - 3,579) | 97 (96 - 98) | 33 (32 - 33) | 32% (32% - 32%) | 11% (11% - 11%) |
| Hawaii | Higher<br>Omicron<br>Severity | 543 (535 - 550) | 233 (227 - 240) | 38 (37 - 38) | 16 (16 - 17) | 39% (39% - 40%) | 17% (16% - 17%) |
| Hawaii | Lower<br>Omicron<br>Severity | 586 (574 - 596) | 188 (179 - 199) | 41 (40 - 41) | 13 (12 - 14) | 42% (41% - 43%) | 14% (13% - 14%) |
| Idaho | Higher<br>Omicron<br>Severity | 2,199 (2,184 - 2,213) | 528 (517 - 540) | 116 (115 - 116) | 28 (27 - 28) | 44% (44% - 44%) | 11% (10% - 11%) |
| Idaho | Lower<br>Omicron<br>Severity | 2,292 (2,279 - 2,305) | 434 (425 - 444) | 121 (120 - 121) | 23 (22 - 23) | 46% (46% - 46%) | 9% (9% - 9%) |
| Illinois | Higher<br>Omicron<br>Severity | 7,228 (7,159 - 7,288) | 4,470 (4,409 - 4,532) | 57 (56 - 58) | 35 (35 - 36) | 21% (21% - 21%) | 13% (13% - 13%) |
| Illinois | Lower<br>Omicron<br>Severity | 8,672 (8,609 - 8,736) | 2,984 (2,939 - 3,036) | 68 (68 - 69) | 24 (23 - 24) | 25% (25% - 25%) | 9% (9% - 9%) |
| Indiana | Higher<br>Omicron<br>Severity | 7,138 (7,069 -<br>7,193) | 2,775 (2,713 - 2,837) | 105 (104 - 106) | 41 (40 - 42) | 30% (30% - 30%) | 12% (11% - 12%) |
| Indiana | Lower<br>Omicron<br>Severity | 7,874 (7,830 -<br>7,914) | 2,029 (1,998 - 2,064) | 116 (115 - 116) | 30 (29 - 30) | 33% (33% - 33%) | 9% (8% - 9%) |
| Iowa | Higher<br>Omicron<br>Severity | 2,486 (2,461 -<br>2,510) | 875 (852 - 896) | 78 (77 - 79) | 27 (27 - 28) | 26% (26% - 26%) | 9% (9% - 9%) |
| Iowa | Lower<br>Omicron<br>Severity | 2,717 (2,698 -<br>2,735) | 645 (631 - 660) | 85 (84 - 86) | 20 (20 - 21) | 28% (28% - 29%) | 7% (7% - 7%) |
| Kansas | Higher<br>Omicron<br>Severity | 2,749 (2,717 -<br>2,778) | 1,102 (1,073 - 1,131) | 94 (93 - 95) | 38 (37 - 39) | 31% (30% - 31%) | 12% (12% - 13%) |
| Kansas | Lower<br>Omicron<br>Severity | 3,044 (3,024 -<br>3,064) | 805 (789 - 824) | 104 (103 - 104) | 27 (27 - 28) | 34% (34% - 34%) | 9% (9% - 9%) |
| Kentucky | Higher<br>Omicron<br>Severity | 6,070 (5,997 - 6,141) | 2,700 (2,635 - 2,763) | 135 (133 - 136) | 60 (58 - 61) | 36% (36% - 37%) | 16% (16% - 17%) |
| Kentucky | Lower<br>Omicron<br>Severity | 6,402 (6,341 - 6,456) | 2,366 (2,329 - 2,413) | 142 (141 - 143) | 52 (52 - 54) | 38% (38% - 39%) | 14% (14% - 15%) |
| Louisiana | Higher<br>Omicron<br>Severity | 4,073 (4,034 - 4,116) | 1,796 (1,768 - 1,820) | 88 (87 - 89) | 39 (38 - 39) | 26% (25% - 26%) | 11% (11% - 11%) |
| Louisiana | Lower<br>Omicron<br>Severity | 4,351 (4,299 - 4,404) | 1,504 (1,468 - 1,549) | 94 (93 - 95) | 33 (32 - 33) | 27% (27% - 28%) | 9% (9% - 10%) |
| Maine | Higher<br>Omicron<br>Severity | 1,025 (1,012 - 1,036) | 419 (406 - 430) | 75 (74 - 76) | 31 (30 - 31) | 41% (41% - 42%) | 17% (16% - 17%) |
| Maine | Lower<br>Omicron<br>Severity | 1,099 (1,089 - 1,109) | 344 (334 - 353) | 80 (79 - 81) | 25 (24 - 26) | 44% (44% - 45%) | 14% (13% - 14%) |
| Maryland | Higher<br>Omicron<br>Severity | 2,824 (2,781 -<br>2,871) | 2,164 (2,116 - 2,207) | 46 (45 - 47) | 35 (34 - 36) | 18% (18% - 19%) | 14% (14% - 14%) |
| Maryland | Lower<br>Omicron<br>Severity | 3,440 (3,377 -<br>3,494) | 1,522 (1,470 - 1,580) | 56 (55 - 57) | 25 (24 - 26) | 22% (22% - 23%) | 10% (9% - 10%) |
| Massachusetts | Higher<br>Omicron<br>Severity | 2,625 (2,595 -<br>2,651) | 1,931 (1,906 - 1,959) | 38 (37 - 38) | 28 (27 - 28) | 14% (14% - 15%) | 11% (10% - 11%) |
| Massachusetts | Lower<br>Omicron<br>Severity | 3,132 (3,109 -<br>3,154) | 1,423 (1,401 - 1,443) | 45 (45 - 45) | 20 (20 - 21) | 17% (17% - 17%) | 8% (8% - 8%) |
| Michigan | Higher<br>Omicron<br>Severity | 9,608 (9,539 -<br>9,670) | 3,546 (3,489 - 3,598) | 96 (95 - 96) | 35 (35 - 36) | 30% (30% - 30%) | 11% (11% - 11%) |
| Michigan | Lower<br>Omicron<br>Severity | 10,609 (10,554 -<br>10,651) | 2,540 (2,501 - 2,578) | 106 (105 - 106) | 25 (25 - 26) | 33% (33% - 33%) | 8% (8% - 8%) |
| Minnesota | Higher<br>Omicron<br>Severity | 3,616 (3,591 -<br>3,640) | 1,350 (1,325 - 1,373) | 63 (63 - 64) | 24 (23 - 24) | 28% (28% - 29%) | 11% (10% - 11%) |
| Minnesota | Lower<br>Omicron<br>Severity | 3,942 (3,923 -<br>3,960) | 1,023 (1,004 - 1,043) | 69 (69 - 69) | 18 (18 - 18) | 31% (31% - 31%) | 8% (8% - 8%) |
| Mississippi | Higher<br>Omicron<br>Severity | 3,473 (3,425 -<br>3,518) | 1,853 (1,815 - 1,908) | 118 (116 - 119) | 63 (62 - 65) | 26% (26% - 27%) | 14% (14% - 14%) |
| Mississippi | Lower<br>Omicron<br>Severity | 3,731 (3,662 -<br>3,800) | 1,582 (1,532 - 1,652) | 126 (124 - 129) | 54 (52 - 56) | 28% (28% - 29%) | 12% (12% - 13%) |
| Missouri | Higher<br>Omicron<br>Severity | 6,551 (6,490 -<br>6,607) | 2,614 (2,565 - 2,669) | 106 (105 - 107) | 42 (42 - 43) | 32% (32% - 32%) | 13% (12% - 13%) |
| Missouri | Lower<br>Omicron<br>Severity | 7,303 (7,255 -<br>7,345) | 1,856 (1,826 - 1,883) | 118 (118 - 119) | 30 (30 - 31) | 36% (35% - 36%) | 9% (9% - 9%) |
| Montana | Higher<br>Omicron<br>Severity | 1,446 (1,430 -<br>1,464) | 351 (333 - 366) | 131 (129 - 133) | 32 (30 - 33) | 41% (41% - 42%) | 10% (9% - 10%) |
| Montana | Lower<br>Omicron<br>Severity | 1,501 (1,488 -<br>1,518) | 295 (281 - 306) | 136 (135 - 137) | 27 (25 - 28) | 43% (42% - 43%) | 8% (8% - 9%) |
| Nebraska | Higher<br>Omicron<br>Severity | 1,386 (1,370 -<br>1,402) | 562 (545 - 578) | 71 (70 - 71) | 29 (28 - 29) | 28% (28% - 28%) | 11% (11% - 12%) |
| Nebraska | Lower<br>Omicron<br>Severity | 1,537 (1,526 -<br>1,549) | 411 (399 - 423) | 78 (78 - 79) | 21 (20 - 22) | 31% (31% - 31%) | 8% (8% - 9%) |
| Nevada | Higher<br>Omicron<br>Severity | 3,555 (3,530 -<br>3,585) | 1,546 (1,521 - 1,568) | 113 (112 - 114) | 49 (48 - 50) | 32% (32% - 33%) | 14% (14% - 14%) |
| Nevada | Lower<br>Omicron<br>Severity | 3,831 (3,807 -<br>3,856) | 1,270 (1,248 - 1,294) | 122 (121 - 123) | 40 (40 - 41) | 35% (35% - 35%) | 12% (11% - 12%) |
| New Hampshire | Higher<br>Omicron<br>Severity | 748 (734 - 764) | 302 (285 - 316) | 54 (53 - 55) | 22 (21 - 23) | 30% (30% - 31%) | 12% (12% - 13%) |
| New Hampshire | Lower<br>Omicron<br>Severity | 828 (818 - 839) | 222 (210 - 232) | 60 (59 - 60) | 16 (15 - 17) | 34% (33% - 34%) | 9% (9% - 9%) |
| New Jersey | Higher<br>Omicron<br>Severity | 3,605 (3,560 -<br>3,654) | 3,501 (3,450 - 3,546) | 39 (38 - 39) | 38 (37 - 38) | 11% (11% - 11%) | 11% (11% - 11%) |
| New Jersey | Lower<br>Omicron<br>Severity | 4,533 (4,453 -<br>4,599) | 2,528 (2,463 - 2,597) | 49 (48 - 50) | 27 (27 - 28) | 14% (14% - 14%) | 8% (8% - 8%) |
| New Mexico | Higher<br>Omicron<br>Severity | 2,583 (2,560 -<br>2,604) | 868 (848 - 890) | 122 (121 - 123) | 41 (40 - 42) | 33% (33% - 34%) | 11% (11% - 11%) |
| New Mexico | Lower<br>Omicron<br>Severity | 2,731 (2,712 -<br>2,751) | 718 (703 - 736) | 129 (128 - 130) | 34 (33 - 35) | 35% (35% - 35%) | 9% (9% - 9%) |
| New York State<br>(non-NYC) | Higher<br>Omicron<br>Severity | 6,096 (6,032 -<br>6,155) | 4,404 (4,348 - 4,469) | 54 (53 - 54) | 39 (38 - 39) | 17% (17% - 17%) | 12% (12% - 12%) |
| New York State<br>(non-NYC) | Lower<br>Omicron<br>Severity | 7,232 (7,163 -<br>7,290) | 3,254 (3,199 - 3,319) | 64 (63 - 64) | 29 (28 - 29) | 20% (20% - 20%) | 9% (9% - 9%) |
| New York City | Higher<br>Omicron<br>Severity | 2,552 (2,508 -<br>2,599) | 2,985 (2,945 - 3,031) | 30 (30 - 31) | 35 (35 - 36) | 7% (7% - 7%) | 9% (8% - 9%) |
| New York City | Lower<br>Omicron<br>Severity | 3,434 (3,376 -<br>3,483) | 2,089 (2,047 - 2,145) | 41 (40 - 41) | 25 (24 - 25) | 10% (10% - 10%) | 6% (6% - 6%) |
| North Carolina | Higher<br>Omicron<br>Severity | 8,522 (8,480 -<br>8,566) | 3,743 (3,705 - 3,782) | 81 (80 - 81) | 35 (35 - 36) | 30% (30% - 30%) | 13% (13% - 13%) |
| North Carolina | Lower<br>Omicron<br>Severity | 9,104 (9,066 -<br>9,144) | 3,152 (3,120 - 3,185) | 86 (86 - 87) | 30 (30 - 30) | 32% (32% - 32%) | 11% (11% - 11%) |
| North Dakota | Higher<br>Omicron<br>Severity | 712 (697 - 728) | 233 (217 - 247) | 92 (90 - 94) | 30 (28 - 32) | 26% (25% - 26%) | 8% (8% - 9%) |
| North Dakota | Lower<br>Omicron<br>Severity | 751 (723 - 762) | 194 (183 - 222) | 97 (93 - 98) | 25 (24 - 29) | 27% (26% - 28%) | 7% (7% - 8%) |
| Ohio | Higher<br>Omicron<br>Severity | 13,800 (13,695 - 13,899) | 6,258 (6,156 - 6,363) | 117 (116 - 118) | 53 (52 - 54) | 32% (32% - 32%) | 14% (14% - 15%) |
| Ohio | Lower<br>Omicron<br>Severity | 15,796 (15,678 - 15,911) | 4,206 (4,116 - 4,299) | 134 (133 - 135) | 36 (35 - 36) | 36% (36% - 37%) | 10% (9% - 10%) |
| Oklahoma | Higher<br>Omicron<br>Severity | 4,863 (4,782 - 4,944) | 2,376 (2,296 - 2,443) | 122 (120 - 124) | 60 (58 - 61) | 30% (30% - 31%) | 15% (14% - 15%) |
| Oklahoma | Lower<br>Omicron<br>Severity | 5,238 (5,178 - 5,288) | 1,995 (1,962 - 2,030) | 131 (130 - 133) | 50 (49 - 51) | 33% (32% - 33%) | 12% (12% - 13%) |
| Oregon | Higher<br>Omicron<br>Severity | 3,074 (3,046 -<br>3,096) | 1,213 (1,193 - 1,236) | 72 (72 - 73) | 29 (28 - 29) | 43% (42% - 43%) | 17% (17% - 17%) |
| Oregon | Lower<br>Omicron<br>Severity | 3,277 (3,251 -<br>3,302) | 1,008 (989 - 1,028) | 77 (77 - 78) | 24 (23 - 24) | 46% (45% - 46%) | 14% (14% - 14%) |
| Pennsylvania | Higher<br>Omicron<br>Severity | 11,937 (11,840 -<br>12,019) | 5,480 (5,398 - 5,572) | 92 (91 - 93) | 42 (42 - 43) | 26% (26% - 26%) | 12% (12% - 12%) |
| Pennsylvania | Lower<br>Omicron<br>Severity | 13,390 (13,307 -<br>13,469) | 4,005 (3,946 - 4,084) | 103 (103 - 104) | 31 (30 - 32) | 29% (29% - 29%) | 9% (9% - 9%) |
| Puerto Rico | Higher<br>Omicron<br>Severity | 788 (770 - 803) | 811 (796 - 829) | 24 (24 - 25) | 25 (24 - 25) | 19% (18% - 19%) | 19% (19% - 20%) |
| Puerto Rico | Lower<br>Omicron<br>Severity | 906 (879 - 927) | 679 (653 - 706) | 28 (27 - 28) | 21 (20 - 22) | 22% (21% - 22%) | 16% (16% - 17%) |
| Rhode Island | Higher<br>Omicron<br>Severity | 487 (475 - 500) | 311 (297 - 323) | 44 (43 - 46) | 28 (27 - 29) | 14% (13% - 14%) | 9% (8% - 9%) |
| Rhode Island | Lower<br>Omicron<br>Severity | 579 (568 - 589) | 219 (209 - 229) | 53 (52 - 54) | 20 (19 - 21) | 16% (16% - 16%) | 6% (6% - 6%) |
| South Carolina | Higher<br>Omicron<br>Severity | 5,378 (5,335 - 5,417) | 2,426 (2,392 - 2,460) | 104 (103 - 104) | 47 (46 - 47) | 30% (30% - 30%) | 14% (13% - 14%) |
| South Carolina | Lower<br>Omicron<br>Severity | 5,711 (5,665 - 5,749) | 2,078 (2,045 - 2,115) | 110 (109 - 111) | 40 (39 - 41) | 32% (32% - 32%) | 12% (11% - 12%) |
| South Dakota | Higher<br>Omicron<br>Severity | 674 (664 - 689) | 265 (251 - 276) | 75 (74 - 77) | 30 (28 - 31) | 22% (22% - 22%) | 9% (8% - 9%) |
| South Dakota | Lower<br>Omicron<br>Severity | 717 (707 - 727) | 221 (212 - 230) | 80 (79 - 81) | 25 (24 - 26) | 23% (23% - 24%) | 7% (7% - 7%) |
| Tennessee | Higher<br>Omicron<br>Severity | 8,581 (8,524 -<br>8,638) | 4,033 (3,979 - 4,088) | 123 (122 - 124) | 58 (57 - 59) | 33% (33% - 33%) | 15% (15% - 16%) |
| Tennessee | Lower<br>Omicron<br>Severity | 9,221 (9,149 -<br>9,274) | 3,380 (3,335 - 3,434) | 132 (131 - 133) | 48 (48 - 49) | 35% (35% - 35%) | 13% (13% - 13%) |
| Texas | Higher<br>Omicron<br>Severity | 26,451 (26,383 -<br>26,529) | 11,854 (11,798 -<br>11,916) | 90 (89 - 90) | 40 (40 - 40) | 28% (28% - 28%) | 12% (12% - 13%) |
| Texas | Lower<br>Omicron<br>Severity | 28,495 (28,383 -<br>28,592) | 9,752 (9,671 - 9,858) | 97 (96 - 97) | 33 (33 - 33) | 30% (30% - 30%) | 10% (10% - 10%) |
| Utah | Higher<br>Omicron<br>Severity | 1,936 (1,917 -<br>1,954) | 609 (595 - 624) | 58 (57 - 59) | 18 (18 - 19) | 38% (37% - 38%) | 12% (12% - 12%) |
| Utah | Lower<br>Omicron<br>Severity | 2,052 (2,035 -<br>2,068) | 485 (474 - 498) | 61 (61 - 62) | 15 (14 - 15) | 40% (40% - 40%) | 9% (9% - 10%) |
| Vermont | Higher<br>Omicron<br>Severity | 164 (158 - 170) | 110 (104 - 116) | 25 (24 - 26) | 17 (16 - 18) | 26% (25% - 27%) | 18% (17% - 18%) |
| Vermont | Lower<br>Omicron<br>Severity | 189 (184 - 195) | 85 (79 - 90) | 29 (28 - 30) | 13 (12 - 14) | 30% (29% - 31%) | 14% (13% - 14%) |
| Virginia | Higher<br>Omicron<br>Severity | 5,063 (5,018 - 5,109) | 3,041 (2,995 - 3,084) | 59 (58 - 59) | 35 (35 - 36) | 26% (26% - 26%) | 16% (15% - 16%) |
| Virginia | Lower<br>Omicron<br>Severity | 5,742 (5,675 - 5,797) | 2,346 (2,297 - 2,414) | 66 (66 - 67) | 27 (27 - 28) | 30% (29% - 30%) | 12% (12% - 12%) |
| Washington | Higher<br>Omicron<br>Severity | 4,262 (4,235 - 4,289) | 1,973 (1,950 - 2,003) | 55 (55 - 55) | 26 (25 - 26) | 35% (35% - 35%) | 16% (16% - 17%) |
| Washington | Lower<br>Omicron<br>Severity | 4,685 (4,649 - 4,714) | 1,551 (1,520 - 1,582) | 61 (60 - 61) | 20 (20 - 20) | 39% (38% - 39%) | 13% (13% - 13%) |
| West Virginia | Higher<br>Omicron<br>Severity | 2,685 (2,655 -<br>2,716) | 1,091 (1,063 - 1,122) | 151 (149 - 152) | 61 (60 - 63) | 39% (39% - 40%) | 16% (16% - 16%) |
| West Virginia | Lower<br>Omicron<br>Severity | 2,865 (2,843 -<br>2,886) | 912 (893 - 932) | 161 (159 - 162) | 51 (50 - 52) | 42% (42% - 42%) | 13% (13% - 14%) |
| Wisconsin | Higher<br>Omicron<br>Severity | 4,505 (4,473 -<br>4,533) | 1,775 (1,749 - 1,803) | 76 (76 - 77) | 30 (30 - 31) | 31% (30% - 31%) | 12% (12% - 12%) |
| Wisconsin | Lower<br>Omicron<br>Severity | 5,058 (5,034 -<br>5,081) | 1,219 (1,199 - 1,243) | 86 (85 - 86) | 21 (20 - 21) | 34% (34% - 35%) | 8% (8% - 8%) |
| Wyoming | Higher<br>Omicron<br>Severity | 749 (736 - 758) | 143 (135 - 155) | 129 (127 - 131) | 25 (23 - 27) | 46% (45% - 47%) | 9% (8% - 10%) |
| Wyoming | Lower<br>Omicron<br>Severity | 767 (756 - 774) | 126 (119 - 136) | 132 (131 - 134) | 22 (21 - 23) | 47% (47% - 48%) | 8% (7% - 8%) |

